# A Longitudinal Psychometric Evaluation of a Context-Sensitive Positive Health Questionnaire for Measuring Broad Health in Dutch Adults

**DOI:** 10.1101/2025.08.23.25334290

**Authors:** Eveline. M. Dubbeldeman, Mirte Boelens, Esther J. Bloemen-van Gurp, John Dierx, Marieke D. Spreeuwenberg, Jessica C. Kiefte-de Jong

**Affiliations:** Leiden University Medical Center/Health Campus The Hague, Department of Public Health and Primary Care, 2511 DP The Hague, The Netherlands; Zuyd University of Applied Sciences, Research Center of community care, Heerlen, The Netherlands; Fontys University of Applied Sciences, Center of Expertise Health, Eindhoven, The Netherlands; Centre of Expertise Perspective in Health, Research Group Equal Chances on Healthy Choices, Avans University of Applied Science, Breda, The Netherlands; Department of Health Services Research, Care and Public Health Research Institute (CAPHRI), Faculty of Health Medicine and Life Sciences, Maastricht University, Maastricht, The Netherlands

**Keywords:** Positive Health, Capability Approach, Factor validity, Concurrent validity, Discriminative validity, Test-retest reliability, Public health, CPHQ

## Abstract

**Introduction:** The Context-sensitive Positive Health Questionnaire (CPHQ) was developed to measure broad health, grounded in Positive Health and the Capability Approach. Through co-creation with professionals from various domains and citizens, including those with lower socioeconomic position, the instrument was revised into a 28-item version. This study evaluates its psychometric properties.

**Methods:** This longitudinal study used a stratified sample of Dutch adults to evaluate the psychometric properties of the revised 28-item CPHQ, including factorial and concurrent validity, reliability, and responsiveness. Factorial validity was assessed using Exploratory Factor Analysis, Item Response Theory, and Confirmatory Factor Analysis. Concurrent validity was examined via correlations with existing health and wellbeing measures. Subgroup comparisons were conducted to assess discriminative validity, using independent samples t-tests or ANOVA. Mixed models and Kappa were used to assess test-retest reliability across three timepoints, controlling for life events.

**Results:** Factor analyses supported an eight-factor structure (mental relaxation, social acceptance, wellbeing, vitality, social support, financial resources, health literacy, mobility) explaining 59.2% of the variance, with good model fit. Internal consistency was acceptable, with composite reliability ranging from 0.59 (health literacy) to 0.90 (financial resources). Concurrent validity was supported by strong correlations with related constructs, particularly in wellbeing, vitality, and social support. Subgroup analyses confirmed hypothesized differences across gender, age, socioeconomic position, and life events, supporting the discriminative validity of the CPHQ2.0. The CPHQ2.0 showed good test-retest reliability (ICCs mostly >0.70), with the total score showing highest stability (ICC=0.88). Kappa values indicated moderate to substantial agreement across timepoints.

**Conclusion:** The CPHQ2.0 is a valid and reliable tool for measuring broad, context-sensitive health, grounded in the Capability Approach and Positive Health concept. Developed with input from diverse stakeholders, it is suitable for use in research, policy, and to target and evaluate health promoting interventions. Future research should assess its sensitivity to detect changes over time following interventions, and explore applicability across settings and populations.

## Introduction

The traditional definition of health, as established by the World Health Organization (1), has often been regarded as insufficient to capture the complexity and multidimensionality of health in modern society. In response, the concept of Positive Health was introduced, defining health as “the ability to adapt and self-manage in the face of social, physical, and emotional challenges” (2). This dynamic and broad perspective contrasts with traditional biomedical models by emphasizing personal strengths, resilience, and the pursuit of a meaningful life, even in the presence of adversity. Rather than focusing solely on disease or limitations, Positive Health focuses on people’s capacity to adapt and self-manage in line with their values, goals, and context (2, 3). This broader view aligns closely with Antonosky’s Salutogenesis approach operationalised in the Sense of Coherence with General and Specific Resistance Resources (4, 5) and the Capability Approach developed by Sen (1993) and Nussbaum (2011). Both approaches highlight not only what people do or achieve (*functionings*), but what they are able to do (*capabilities*), given their personal and contextual circumstances. Together, these frameworks move beyond viewing health as a static or clinical condition, offering instead a dynamic lens that accounts for real-life complexity, individual agency, and structural context. By recognizing health as a context-sensitive and evolving phenomenon shaped by social, economic, and environmental determinants, Positive Health and the Capability Approach enable a more inclusive and meaningful understanding of health—also for people in vulnerable or disadvantaged positions (3, 8).

Several attempts have been made to operationalize this broader perspective on health. One of the first was a 44-item Positive Health questionnaire, which was criticized for lacking comprehensiveness and discriminant validity (9). A shortened 17-item version showed improved psychometric properties (10), but failed to capture essential elements such as empowerment and contextual factors like financial security and living conditions (11). These elements are central to the Capability Approach, which emphasizes the importance of individual resources and opportunities for achieving well-being (6, 7). To better integrate the broad and dynamic nature of Positive Health with the Capability Approach’s focus on these resources and opportunities, the Context-sensitive Positive Health Questionnaire (CPHQ) was developed (11). This 32-item questionnaire showed promising psychometric properties, but also had some limitations. Although grounded in both Positive Health and the Capability Approach, the alignment between the theoretical framework and the item structure was only partial. Conceptual domains such as social support, exclusion, and environmental safety were included but showed weak empirical associations with existing validation measures, raising concerns about their integration and measurement relevance. Additionally, stakeholder involvement during development was limited, particularly from professionals in the medical, social, and policy domains—whose perspectives are essential for broad applicability of the CPHQ. Finally, there was insufficient evidence regarding the instrument’s applicability to diverse and vulnerable populations, such as individuals in vulnerable or disadvantaged positions (11, 12).

To address these issues, the original CPHQ was revised in co-creation with stakeholders from healthcare, welfare, and policy sectors, as well as with direct input from citizens and patients, including those with a lower socioeconomic position (SEP) (12). Through structured focus group discussions, stakeholders, citizens and patients critically assessed the original items for comprehensibility, relevance, and cultural sensitivity. They also identified domains that were insufficiently covered in the initial questionnaire. Consequently, several items were reformulated to improve clarity and to better capture contextual nuances, with a stronger emphasis on social inclusion, meaningfulness, and resilience. This iterative, participatory process resulted in a more concise 28-item version that is better aligned with the perspectives of both professionals and end-users. Although this revised version better reflects the lived experiences of diverse users, systematic quantitative validation is necessary to establish its psychometric quality. Such validation is essential to confirm reliability, dimensional structure, and generalizability across different populations, ensuring the instrument accurately and consistently measures the intended constructs.

The aim of this study is to evaluate the psychometric properties of the revised 28-item CPHQ (12). Specifically, we will assess its factorial validity, concurrent validity, and responsiveness, including test-retest reliability and discriminative validity (its ability to detect relevant differences between subgroups).

## Methods

### Study Design

This study employed a longitudinal design (T0, T1, T2), with two-week intervals between each measurement. Data was collected through an online questionnaire in collaboration with Flycatcher, a research agency based in Maastricht, the Netherlands (13). The Medical Ethics Committee at Leiden University Medical Center (LUMC) [Proposal Number 19035] approved the study. Reporting follows the STROBE guidelines (14).

### Participants and Procedures

Participants were recruited through the Dutch Flycatcher’s online research panel. The target population consisted of residents of the Netherlands aged 18 years and older. A stratified sampling method was used to select participants based on gender, age, SEP, and region, ensuring that the sample was representative of the Dutch population. Flycatcher’s panel consists of over 10,000 individuals who have voluntarily opted in to participate in online research. Panel members cannot choose the types of studies for which they receive invitations.

Selected participants received an email invitation containing a unique hyperlink to access the questionnaire. To ensure data quality, participants first answered a verification question to prevent other individuals, such as household members, from completing the questionnaire on their behalf. Access to the questionnaire was restricted to invited participants only, enhancing data reliability. Panel members received a fixed number of reward points for completing the questionnaire, which they could later exchange for gift vouchers, along with an entry into Flycatcher’s lottery. Participation was entirely voluntary, and panel members could withdraw from the panel at any time. Before starting the study, all participants were informed about the purpose and procedures, and they provided informed consent electronically before accessing the questionnaire.

We aimed to collect data from at least 1,000 participants, which—based on recommendations by Comrey and Lee (2013) and Worthington and Whittaker (2006)—provides an adequate sample size and a participant-to-item ratio of approximately 8.5:1, suitable for conducting factor analysis.

### Data Collection

The questionnaire for T0 was distributed to 1,740 panel members on February 16th, 2024, with a response deadline of February 26th, 2024. A reminder email was sent on February 20th to participants who had not yet completed the questionnaire, and an additional group of participants was invited on February 21st to boost response rates. For T1, the questionnaire was distributed between March 8th and March 15th, 2024, to a subsample of 870 panel members. For T2, the questionnaire was distributed between March 29th and April 5th, 2024, to 623 panel members. All data were collected and processed according to ISO 27001 and ISO 20252, ensuring secure storage of information, participant anonymity through data categorization and anonymization, and the integrity of research processes by adhering to rigorous quality standards.

### Questionnaire

The survey was developed by researchers from Maastricht University, Leiden University Medical Center, and Zuyd University of Applied Science in consultation with Flycatcher. The questionnaire was extensively pretested internally by two researchers (other than the principal investigator) for both content and technical aspects. Flycatcher also provided researchers the opportunity to review and navigate the digital questionnaire via a test account before its distribution. The final version of the questionnaire was programmed by Flycatcher, ensuring the technical functionality of the online format and implementing quality checks.

At each timepoint, participants completed a survey. At T0, participants completed the revised 28-item CPHQ (12). Each item was rated on a 5-point Likert scale, where higher scores reflected more positive responses and lower scores more negative ones. One item was negatively worded, for which the scoring was reversed, with higher scores indicating more negative responses. The survey also included items related to positive and adverse life events experienced in the past year (e.g., marriage, birth, accidents), work situation (e.g., retired, disabled, unemployed), and non-communicable diseases (e.g., diabetes, neck and back pain, cancer). In addition, several validated instruments were administered: the EuroQol 5 Dimensions 5 Levels (EQ5D-5L) for health-related quality of life (17), the Brief Resilience Scale (BRS) which evaluates an individual’s ability to recover from stress (18), the ICEpop CAPability measure for Adults (ICECAP-A) measures capability wellbeing (19), the Positive Health 17-item measure (PH-17) examines positive health (10), and the Personal Wellbeing Index (PWI) for psychological wellbeing (20). Demographic data (e.g., gender, age, education, socioeconomic position, province, region, income, living situation, and country of birth) were provided by Flycatcher and were not part of the survey. At T1 and T2, participants again completed the revised 28-item CPHQ, along with items assessing positive and adverse life events that occurred in the past two weeks.

### Analyses

To evaluate the factorial validity of the revised 28-item CPHQ — which included both modified and newly developed items — we conducted Exploratory Factor Analysis (EFA), Item Response Theory (IRT), and Confirmatory Factor Analysis (CFA). Analyses were performed using RStudio, version 2023.06.1. To enhance the validity of our findings, the dataset was randomly divided into two subsets. We examined potential significant differences in socio-demographic characteristics between these subsets and found none (Appendix A). The first subset was used for EFA and IRT, while the second subset was reserved for CFA.

#### Factorial validity

EFA was conducted to explore the underlying factor structure of 28 items. Data suitability for factor analysis was confirmed using the Kaiser-Meyer-Olkin (KMO) measure of sampling adequacy (>0.7) and Bartlett’s test of sphericity (p<0.05), indicating that the data met the assumptions for EFA. The analysis was performed using the ‘psych’ package, employing the weighted least squares estimation method, which is suitable for ordinal Likert-type data. Oblimin rotation was applied to account for potential correlations between factors, acknowledging the multidimensional aspects of health (21). The number of factors to retain was determined by parallel analysis, total variance explained, and evaluating fit indices (i.e., Comparative Fit Index (CFI), Tucker-Lewis Index (TLI), Root Mean Square Error of Approximation (RMSEA), and Root Mean Square Residual (RMSR) (22)). Additionally, the interpretability of the factors was considered. Items were retained if they demonstrated strong loadings (≥0.40) on their primary factor, with no cross-loadings exceeding this threshold.

Following the EFA, an IRT analysis was performed to assess the quality of individual items within factors. A Generalized Partial Credit Model, suitable for ordinal data, was employed using the ‘mirt’ package (23). Items were retained if they had strong loadings (≥0.70), communalities (≥0.40), and item information values (>1.0), ensuring a strong relationship with the latent factors and sufficient measurement precision across the factor spectrum.

We conducted a CFA using the ‘lavaan’ package to validate the factor structure of the CPHQ2.0 questionnaire. The Diagonally Weighted Least Squares estimator, suitable for ordinal data, was employed (24). Model fit was assessed using CFI and TLI (acceptable >0.95) and RMSEA (acceptable <0.06) and RMSR (acceptable <0.08) (25). Items with standardized factor loadings (λ ≥0.70) and R² values >0.40 were retained, confirming that the latent constructs explained a substantial portion of the variance in the observed variables and supporting the model’s factorial validity.

Composite Reliability (CR) was calculated to assess internal consistency, ensuring that the items within each construct reliably measure the same underlying factor, with a CR value above 0.60 indicating acceptable reliability. Similarly, Average Variance Extracted (AVE) was used to assess convergent validity, indicating the extent to which the items explain the variance in the construct. An AVE value above 0.50 suggests that the items are adequately related to the construct, confirming its validity (26).

#### CPHQ2.0 scoring

The interpretation of the CPHQ2.0 is based on domain scores and a total score. These were derived through a stepwise process. First, the negatively worded item was reverse-coded. Next, all item scores were rescaled to a 0–100 scale, with higher values indicating better health. Domain scores were then calculated as the average of all item scores within each respective domain. Finally, the total CPHQ2.0 score was computed as the average of all domain scores.

#### Concurrent validity

To evaluate the relationship between the CPHQ and related constructs, we conducted correlation analyses with the previously described validation instruments (EQ5D-5L, BRS, ICECAP-A, PH-17, and PWI). Correlation coefficients below 0.30 were interpreted as indicating weak validity, values between 0.30 and 0.70 as moderate validity, and values above 0.70 as strong validity.

#### Discriminative validity

Subgroup analyses were conducted using independent samples t-tests or ANOVA, including post-hoc tests where necessary, to examine discriminative validity (i.e., differences in CPHQ outcomes across various subgroups). These analyses were based on hypotheses regarding differences in CPHQ scores related to gender, age, migration background, education, income, SEP, household composition, adverse and positive events, and non-communicable diseases (Appendix B). For example, we hypothesized that participants with higher income will score higher (indicating better health) on the CPHQ2.0 compared to those with lower income (27), and that participants with non-communicable diseases would score lower (indicating poorer health) than those without non-communicable diseases (28).

#### Test-retest reliability

Test-retest reliability of the CPHQ2.0 domain scores and total score was assessed using linear mixed models and weighted Kappa analysis.

Linear mixed models examined the effects of time (T0, T1, T2) on scores, while controlling for life events (adverse and positive events at each time point). Time and event types were included as fixed effects, with a random intercept to account for individual variability. An autoregressive covariance structure was applied to model within-subject correlations across time. Models were estimated using restricted maximum likelihood, and Satterthwaite’s method was used to calculate degrees of freedom. Intraclass Correlation Coefficients (ICCs) were calculated to assess test-retest reliability.

For the weighted Kappa analyses, standardized residuals from linear regression models were used to determine agreement between scores at each time point. The regression models adjusted for the impact of positive and adverse events on scores over time. Residuals were categorized into four levels based on standardized score ranges at each time point: category 1=residuals<-1, category 2=residuals -1 to<0, category 3=residuals 0 to<1, and category 4=residuals ≥ 1. We applied quadratic weighting to account for the severity of discrepancies between adjacent categories of residuals. The weighted Kappa coefficients and 95% confidence intervals were then calculated to determine the level of agreement between time points, with coefficients >0.8 considered as excellent, 0.61–0.8 as substantial, 0.41–0.60 as moderate, 0.21–0.40 as fair, and 0.01–0.2 as slight agreement (29).

## Results

Of the 1,740 panel members sampled at T0, 1,738 surveys were successfully distributed. After excluding 31 cases due to incomplete responses or poor response quality, a total of 1,043 valid responses were received, yielding a 60% response rate. Of these, 1,001 participants provided consent for research use. At T1, 870 panel members were sampled, of which 869 surveys were successfully distributed. Eight responses were excluded due to incomplete or poor quality, resulting in 714 valid responses (82% response rate). At T2, surveys were distributed to 623 panel members, all successfully delivered. After excluding five responses for quality reasons, 543 valid responses were obtained (87% response rate). Respondents were representative of the Dutch population aged 18 and older in terms of gender, SEP, and region of residence. However, at T0. individuals aged 60 and older were overrepresented (Appendix A).

### Factorial validity

Data suitability for factor analysis was confirmed (KMO=0.924, Bartlett’s χ²(378)=7642.67, p<.001). Parallel analysis and EFA revealed an eight-factor solution, accounting for 59.2% of the total variance (Appendix A). The factor loadings ranged from 0.430 to 1.02, indicating a strong representation of items on their respective factors (Table 1). Fit indices indicated good model fit (CFI=0.97, TLI=0.95, RMSEA=0.05, RSMR=0.02).

**Table 1.**
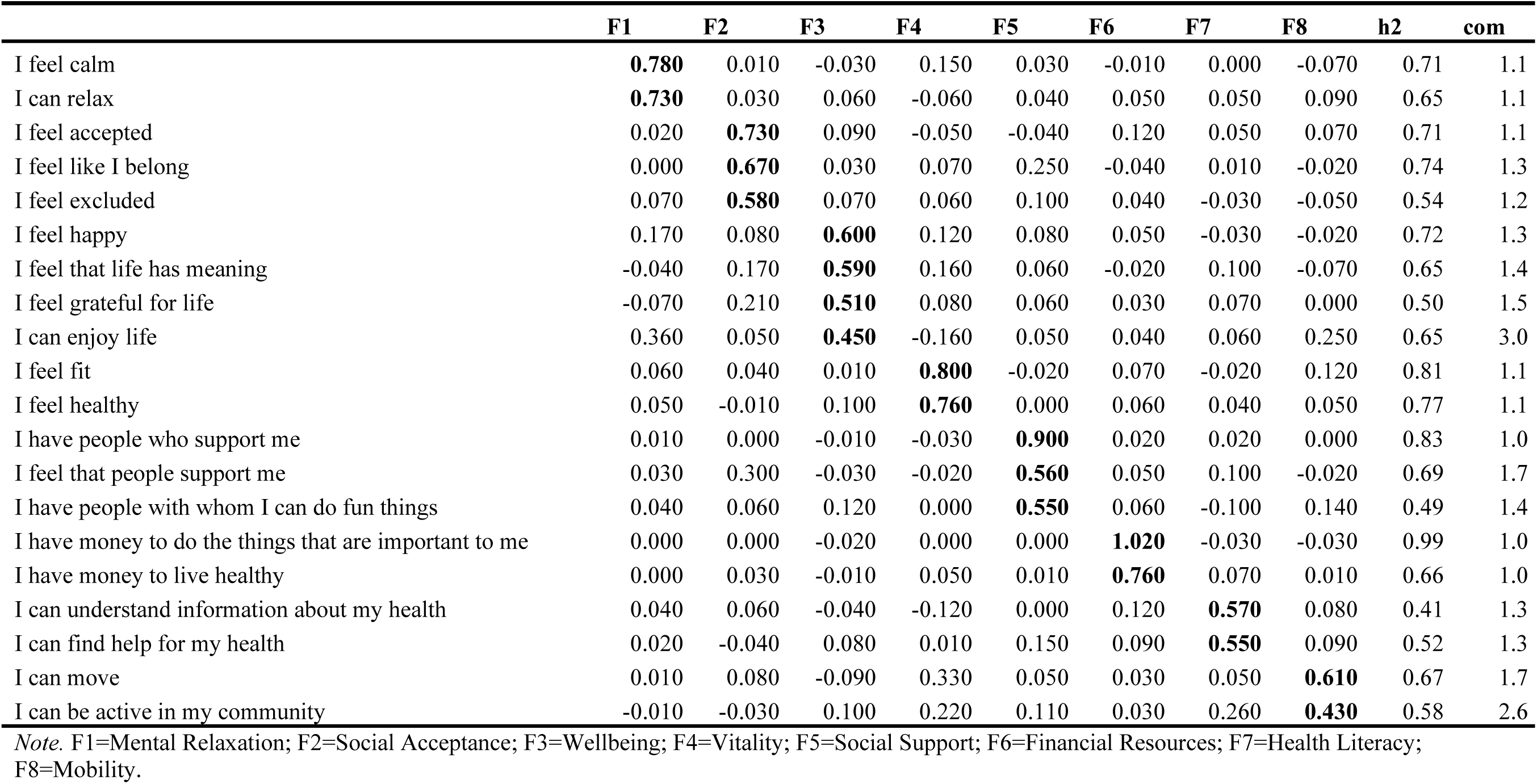
Standardized weights of the exploratory eight-factor model using oblimin rotated factors.

|  | F1 | F2 | F3 | F4 | F5 | F6 | F7 | F8 | h2 | com |
| --- | --- | --- | --- | --- | --- | --- | --- | --- | --- | --- |
| I feel calm | **0.780** | 0.010 | -0.030 | 0.150 | 0.030 | -0.010 | 0.000 | -0.070 | 0.71 | 1.1 |
| I can relax | **0.730** | 0.030 | 0.060 | -0.060 | 0.040 | 0.050 | 0.050 | 0.090 | 0.65 | 1.1 |
| I feel accepted | 0.020 | **0.730** | 0.090 | -0.050 | -0.040 | 0.120 | 0.050 | 0.070 | 0.71 | 1.1 |
| I feel like I belong | 0.000 | **0.670** | 0.030 | 0.070 | 0.250 | -0.040 | 0.010 | -0.020 | 0.74 | 1.3 |
| I feel excluded | 0.070 | **0.580** | 0.070 | 0.060 | 0.100 | 0.040 | -0.030 | -0.050 | 0.54 | 1.2 |
| I feel happy | 0.170 | 0.080 | **0.600** | 0.120 | 0.080 | 0.050 | -0.030 | -0.020 | 0.72 | 1.3 |
| I feel that life has meaning | -0.040 | 0.170 | **0.590** | 0.160 | 0.060 | -0.020 | 0.100 | -0.070 | 0.65 | 1.4 |
| I feel grateful for life | -0.070 | 0.210 | **0.510** | 0.080 | 0.060 | 0.030 | 0.070 | 0.000 | 0.50 | 1.5 |
| I can enjoy life | 0.360 | 0.050 | **0.450** | -0.160 | 0.050 | 0.040 | 0.060 | 0.250 | 0.65 | 3.0 |
| I feel fit | 0.060 | 0.040 | 0.010 | **0.800** | -0.020 | 0.070 | -0.020 | 0.120 | 0.81 | 1.1 |
| I feel healthy | 0.050 | -0.010 | 0.100 | **0.760** | 0.000 | 0.060 | 0.040 | 0.050 | 0.77 | 1.1 |
| I have people who support me | 0.010 | 0.000 | -0.010 | -0.030 | **0.900** | 0.020 | 0.020 | 0.000 | 0.83 | 1.0 |
| I feel that people support me | 0.030 | 0.300 | -0.030 | -0.020 | **0.560** | 0.050 | 0.100 | -0.020 | 0.69 | 1.7 |
| I have people with whom I can do fun things | 0.040 | 0.060 | 0.120 | 0.000 | **0.550** | 0.060 | -0.100 | 0.140 | 0.49 | 1.4 |
| I have money to do the things that are important to me | 0.000 | 0.000 | -0.020 | 0.000 | 0.000 | **1.020** | -0.030 | -0.030 | 0.99 | 1.0 |
| I have money to live healthy | 0.000 | 0.030 | -0.010 | 0.050 | 0.010 | **0.760** | 0.070 | 0.010 | 0.66 | 1.0 |
| I can understand information about my health | 0.040 | 0.060 | -0.040 | -0.120 | 0.000 | 0.120 | **0.570** | 0.080 | 0.41 | 1.3 |
| I can find help for my health | 0.020 | -0.040 | 0.080 | 0.010 | 0.150 | 0.090 | **0.550** | 0.090 | 0.52 | 1.3 |
| I can move | 0.010 | 0.080 | -0.090 | 0.330 | 0.050 | 0.030 | 0.050 | **0.610** | 0.67 | 1.7 |
| I can be active in my community | -0.010 | -0.030 | 0.100 | 0.220 | 0.110 | 0.030 | 0.260 | **0.430** | 0.58 | 2.6 |

Note. F1=Mental Relaxation; F2=Social Acceptance; F3=Wellbeing; F4=Vitality; F5=Social Support; F6=Financial Resources; F7=Health Literacy; F8=Mobility.

Overall, the IRT analysis showed strong item loadings (0.77 to 0.99), indicating good model fit and substantial relationships with the latent constructs. Item communalities were consistently above 0.5, reflecting reliable measurement. Most items exhibited high discrimination (a-parameters), effectively differentiating individuals across the traits. The difficulty (b-parameters) varied, contributing to an appropriate spread of item difficulty, and item information values were mostly above 1.0, ensuring sufficient measurement precision across the factors (Appendix C).

The CFA results indicate strong model fit, showing excellent performance considering the CFI (0.997), TLI (0.996), RMSEA (0.046), and SRMR (0.044). Factor loadings were generally high, with values between 0.678 and 0.993, and R² values ranged from 0.460 to 0.987, indicating adequate construct validity and item reliability (Table 2). Covariances were positive, reflecting meaningful relationships among factors. Financial Resources exhibited the highest reliability (CR = 0904, AVE = 0.919), indicating strong internal consistency and convergent validity. In contrast, Health Literacy showed the lowest values (CR = 0.593, AVE = 0.567), suggesting weaker performance (Table 3).

**Table 2.** Parameter estimates Confirmatory Factor Analysis.

| Latent Factor | Item | B | SE | Z | $\beta$ | R<sup>2</sup> | p |
| --- | --- | --- | --- | --- | --- | --- | --- |
| Mental Relaxation | I feel calm | 1.000 | 0.020 | 42.857 | 0.855 | 0.731 |  |
| Mental Relaxation | I can relax | 1.002 | 0.020 | 42.857 | 0.856 | 0.733 | *** |
| Social Acceptance | I feel accepted | 1.000 | 0.013 | 69.616 | 0.917 | 0.841 |  |
| Social Acceptance | I feel that I belong | 0.997 | 0.013 | 70.091 | 0.915 | 0.837 | *** |
| Social Acceptance | I feel excluded* | 0.869 | 0.014 | 58.039 | 0.797 | 0.635 | *** |
| Wellbeing | I feel happy | 1.000 | 0.013 | 70.728 | 0.902 | 0.813 |  |
| Wellbeing | I feel my life is meaningful | 0.879 | 0.013 | 61.416 | 0.793 | 0.628 | *** |
| Wellbeing | I feel grateful for my life | 0.891 | 0.013 | 64.181 | 0.803 | 0.645 | *** |
| Wellbeing | I can enjoy | 0.951 | 0.014 | 61.276 | 0.858 | 0.736 | *** |
| Vitality | I feel fit | 1.000 | 0.015 | 58.391 | 0.881 | 0.775 |  |
| Vitality | I feel healthy | 1.084 | 0.016 | 58.391 | 0.955 | 0.911 | *** |
| Social Support | I have people who support me | 1.000 | 0.013 | 67.535 | 0.874 | 0.763 |  |
| Social Support | I feel that people support me | 1.083 | 0.014 | 68.670 | 0.946 | 0.895 | *** |
| Social Support | I have people with I can do fun things with | 0.942 | 0.014 | 57.119 | 0.823 | 0.678 | *** |
| Financial Resources | I have money to do the things that I find important | 1.000 | 0.021 | 47.834 | 0.993 | 0.987 |  |
| Financial Resources | I have money to live a healthy life | 0.929 | 0.019 | 47.834 | 0.922 | 0.851 | *** |
| Health Literacy | I can understand information about my health | 1.000 | 0.026 | 26.009 | 0.678 | 0.460 |  |
| Health Literacy | I can find help for my health | 1.210 | 0.032 | 26.009 | 0.821 | 0.674 | *** |
| Mobility | I can move | 1.000 | 0.024 | 31.368 | 0.751 | 0.565 |  |
| Mobility | I can be active in my environment | 1.128 | 0.027 | 31.368 | 0.848 | 0.718 | *** |

Note. \* Negative formulation; B=unstandardized factor loadings; $\beta$ =standardized factor loadings; R<sup>2</sup>=explained variance.

**Table 3.** Correlations among the factors based on the test dataset.

|  | **Mental**<br>**Relaxation** | **Social**<br>**Acceptance** | **Wellbeing** | **Vitality** | **Social**<br>**Support** | **Financial**<br>**Resources** | **Health**<br>**Literacy** | **Mobility** |
| --- | --- | --- | --- | --- | --- | --- | --- | --- |
| **Mental Relaxation** | (0.759, 0.732) |  |  |  |  |  |  |  |
| **Social Acceptance** | 0.45*** | (0.832, 0.771) |  |  |  |  |  |  |
| **Wellbeing** | 0.56*** | 0.64*** | (0.850, 0.705) |  |  |  |  |  |
| **Vitality** | 0.44*** | 0.38*** | 0.53*** | (0.848, 0.843) |  |  |  |  |
| **Social Support** | 0.33*** | 0.68*** | 0.60*** | 0.35*** | (0.837, 0.779) |  |  |  |
| **Financial Resources** | 0.22*** | 0.38*** | 0.37*** | 0.37*** | 0.40*** | (0.904, 0.919) |  |  |
| **Health Literacy** | 0.27*** | 0.41*** | 0.39*** | 0.35*** | 0.40*** | 0.43*** | (0.593, 0.567) |  |
| **Mobility** | 0.33*** | 0.40*** | 0.46*** | 0.62*** | 0.44*** | 0.41*** | 0.46*** | (0.677, 0.641) |

*Note.* Values in brackets: Composite Reliability, Average Variance Extracted.

### Concurrent validity

The validity of the CPHQ2.0 was supported by the significant correlations with various validation scales (Table 4). Strong correlations were primarily observed in the CPHQ 2.0 domains wellbeing, vitality, and the total score. Wellbeing showed strong correlations with PH-17 contentment (r=0.76), PWI life satisfaction (r=0.74), and PWI total score (r=0.74). The vitality domain was strongly correlated with PH-17 physical fitness (r=0.82), PH-17 total score (r=0.72), and PWI health (r=0.81). The total score demonstrated strong correlations with PH-17 contentment (r=0.74), PH-17 total score (r=0.78), and PWI total score (r=0.80). Additionally, a strong correlation was found between the CPHQ2.0 social support domain and PH-17 social relations (r=0.72). Moderate correlations were found in the remaining CPHQ2.0 domains. Mental relaxation had the highest correlations with PH-17 contentment (r=0.59), EQ5D-5L anxiety/depression (r=-0.55), and PH-17 total score (r=-0.54). Mobility showed the strongest correlations with PH-17 physical fitness (r=0.65), PH-17 total score (r=0.58), and PWI health (r=0.58). The strongest correlations for social acceptance were observed with PH-17 social relations (r=0.68), PWI community-connectedness (r=0.63), and PWI total score (r=0.62). Financial resources had a relatively strong correlation with PWI standard of living (r=0.60). The CPHQ2.0 health literacy domain was the least strongly correlated with the validation scales, with the strongest correlation being with PH-17 total score (r=0.41).

**Table 4.** Correlation matrix between the eight factors and validation scales.

| Scale | Domain | Mental<br>Relaxation | Social<br>Acceptance | Wellbeing | Vitality | Social<br>Support | Financial<br>Resources | Health<br>Literacy | Mobility | Total<br>score |
| --- | --- | --- | --- | --- | --- | --- | --- | --- | --- | --- |
| EQ5D-5L | Mobility | -0.08* | -0.13*** | -0.19*** | -0.45*** | -0.17*** | -0.22*** | -0.19*** | -0.47*** | -0.34*** |
|  | Self-care | -0.05 | -0.08* | -0.09** | -0.29*** | -0.06 | -0.11*** | -0.09** | -0.28*** | -0.19*** |
|  | Usual activities | -0.22*** | -0.27*** | -0.32*** | -0.58*** | -0.19*** | -0.29*** | -0.23*** | -0.49*** | -0.47*** |
|  | Pain/Discomfort | -0.19*** | -0.19*** | -0.26*** | -0.55*** | -0.20*** | -0.27*** | -0.19*** | -0.42*** | -0.41*** |
|  | Anxiety/Depression | -0.55*** | -0.44*** | -0.51*** | -0.33*** | -0.31*** | -0.14*** | -0.16*** | -0.20*** | -0.47*** |
|  | EQ-VAS | 0.36*** | 0.32*** | 0.43*** | 0.66*** | 0.25*** | 0.29*** | 0.24*** | 0.47*** | 0.55*** |
|  | EQ5D-5L index | 0.38*** | 0.36*** | 0.44*** | 0.61*** | 0.30*** | 0.29*** | 0.25*** | 0.50*** | 0.56*** |
| BRS | Resilience | 0.48*** | 0.39*** | 0.42*** | 0.32*** | 0.26*** | 0.20*** | 0.27*** | 0.23*** | 0.45*** |
| ICECAP-A | Stability | -0.39*** | -0.40*** | -0.43*** | -0.33*** | -0.28*** | -0.22*** | -0.21*** | -0.28*** | -0.45*** |
|  | Attachment | -0.34*** | -0.49*** | -0.52*** | -0.28*** | -0.56*** | -0.21*** | -0.25*** | -0.32*** | -0.52*** |
|  | Mobility | -0.29*** | -0.31*** | -0.31*** | -0.34*** | -0.23*** | -0.26*** | -0.27*** | -0.30*** | -0.41*** |
|  | Achievement | -0.28*** | -0.32*** | -0.42*** | -0.48*** | -0.33*** | -0.32*** | -0.26*** | -0.45*** | -0.51*** |
|  | Wellbeing | -0.45*** | -0.43*** | -0.61*** | -0.41*** | -0.46*** | -0.27*** | -0.27*** | -0.41*** | -0.58*** |
|  | Total score | 0.48*** | 0.56*** | 0.66*** | 0.50*** | 0.57*** | 0.36*** | 0.34*** | 0.50*** | 0.70*** |
| PH-17 | Physical fitness | 0.33*** | 0.32*** | 0.45*** | 0.82*** | 0.30*** | 0.34*** | 0.29*** | 0.65*** | 0.64*** |
|  | Mental functioning | 0.42*** | 0.34*** | 0.40*** | 0.43*** | 0.29*** | 0.25*** | 0.28*** | 0.28*** | 0.48*** |
|  | Future perspectives | 0.36*** | 0.40*** | 0.59*** | 0.55*** | 0.42*** | 0.32*** | 0.36*** | 0.43*** | 0.61*** |
|  | Contentment | 0.59*** | 0.54*** | 0.76*** | 0.64*** | 0.48*** | 0.32*** | 0.31*** | 0.48*** | 0.74*** |
|  | Social relations | 0.43*** | 0.68*** | 0.66*** | 0.39*** | 0.72*** | 0.30*** | 0.37*** | 0.43*** | 0.69*** |
|  | Daily life-management | 0.43*** | 0.36*** | 0.42*** | 0.48*** | 0.30*** | 0.27*** | 0.36*** | 0.41*** | 0.54*** |
|  | Total score | 0.54*** | 0.55*** | 0.69*** | 0.72*** | 0.52*** | 0.38*** | 0.41*** | 0.58*** | 0.78*** |
| PWI | Life satisfaction | 0.48*** | 0.51*** | 0.74*** | 0.57*** | 0.48*** | 0.35*** | 0.31*** | 0.44*** | 0.69*** |
|  | Standard of living | 0.37*** | 0.44*** | 0.55*** | 0.47*** | 0.42*** | 0.60*** | 0.37*** | 0.43*** | 0.65*** |
|  | Health | 0.40*** | 0.36*** | 0.52*** | 0.81*** | 0.33*** | 0.34*** | 0.31*** | 0.58*** | 0.66*** |
|  | Achieving in life | 0.43*** | 0.51*** | 0.65*** | 0.52*** | 0.44*** | 0.37*** | 0.29*** | 0.39*** | 0.64*** |
|  | Relationships | 0.42*** | 0.56*** | 0.65*** | 0.38*** | 0.60*** | 0.30*** | 0.27*** | 0.35*** | 0.62*** |
|  | Safety | 0.36*** | 0.39*** | 0.47*** | 0.35*** | 0.36*** | 0.30*** | 0.30*** | 0.37*** | 0.51*** |
|  | Community-connectedness | 0.43*** | 0.63*** | 0.62*** | 0.42*** | 0.55*** | 0.29*** | 0.34*** | 0.40*** | 0.65*** |
|  | Future security | 0.44*** | 0.51*** | 0.61*** | 0.54*** | 0.45*** | 0.40*** | 0.35*** | 0.44*** | 0.67*** |
|  | Total score | 0.52*** | 0.62*** | 0.74*** | 0.64*** | 0.57*** | 0.46*** | 0.40*** | 0.54*** | 0.80*** |

*Note.* EQ5D-5L = EuroQol 5-Dimensional; EQ-VAS = EuroQol Visual Analog Scale; BRS = Brief Resilience Scale; ICECAP-A = ICEpop CAPability measure for Adults; PH-17=Positive Health 17 item; PWI=Personal Wellbeing Index.

### Discriminative validity

The findings confirm most of the hypotheses (Appendix A) and support the discriminative validity of the CPHQ2.0. Men generally had higher scores across the CPHQ domains and total score than women, with a significant difference in *mental relaxation* (mean: 73.1 vs. 68.2, *p*<0.001), social acceptance (mean: 78.3 vs. 74.5, *p*<0.001), and total score (mean: 76.1 vs. 74.7, *p*=0.045). Participants younger than 65 years scored significantly higher on most domains, except for *mental relaxation and social acceptance*, where they scored significant lower (mean: 69.4 vs. 73.9, *p*<0.001 *and* 75.7 vs. 78.1, *p*<0.014, respectively). Participants with both parents born in the Netherlands scored higher on all domains and total score, with significant differences observed in *financial resources* (mean: 80.4 vs. 75.4, *p*=0.010), *health literacy* (mean: 81.7 vs. 77.7, *p*=0.011), mobility (mean: 77.5 vs. 74.2, *p*=0.040), and total score (mean: 75.7 vs. 72.2, *p*=0.007). Participants with lower income, educational level, and SEP scored significantly lower across all domains (*p*<0.001), except for *mental relaxation*, where the difference was not significant. Regarding household composition, participants living without a partner scored lower than those with a partner, with significant differences in social acceptance (mean: 72.5 vs. 77.8, *p*<0.001), *wellbeing* (mean: 70.9 vs. 76.3, *p*<0.001), *social support* (mean: 73.4 vs. 77.2, *p*<0.001), *financial resources* (mean: 76.0 vs. 81.5, *p*<0.001), and total score (mean: 72.9 vs. 76.3, *p*<0.001). We did not account for the presence of children in the household. Participants who experienced adverse events had lower scores that those who did not, with significant differences in *mental relaxation* (mean: 67.2 vs. 73.2, *p*<0.001), social acceptance (mean: 75.3 vs. 77.2, *p*=0.047), *wellbeing* (mean: 73.3 vs. 75.9, *p*=0.007), *mobility* (mean: 75.6 vs. 78.3, *p*=0.004), and total score (mean: 73.9 vs. 76.5, *p*<0.001). A higher number of adverse events further decreased scores, with an additional significant difference in the *vitality* domain (mean: none=69.7; one=64.2; two or more=59.8, *p<*0.001). Positive events resulted in significant higher scores in social support (mean: 78.0 vs. 75.5, p=0.017), health literacy (mean: 82.7 vs. 80.9 p=0.044, and mobility (mean: 78.8 vs. 76.6, p=0.038). Significant differences were observed in social support (mean: none=75.5; one=77.1; two or more=78.5, *p=*0.040) and health literacy (mean: none=80.9; one=80.6; two or more=83.9, *p=*0.019) with increasing numbers of positive events. Lastly, the presence of non-communicable diseases was associated with significant lower scores across all domains and total score.

### Test-retest reliability

The mixed model analysis showed that time had generally no significant impact on scores across domains and total score, indicating that the scores remained largely stable over time (Appendix A). An exception was observed in social acceptance, where a slight decrease in score was noted between T0 and T1 (β=- 0.844), p=0.043). Adverse life events had a significant impact on several domains. Lower scores (indicating deterioration) were observed in mental relaxation, wellbeing, vitality, and total score following adverse events. In contrast, positive events did not lead to significant improvements in scores in any domain or total score. T

The ICCs were generally high (mostly >0.70), indicating good test-retest reliability of the CPHQ2.0 scores. The total score demonstrated the highest stability (ICC=0.877), following wellbeing (ICC=0.848), social acceptance (ICC=0.784), vitality (ICC=0.777), financial resources (ICC=0.752), mental relaxation (ICC=0.751), social support (ICC=0.733), and mobility (ICC=0.718). The health literacy domain showed a slightly lower ICC (0.617), indicating greater variability in measurements within this domain.

Weighted Kappa coefficients for T0 and T1 ranged from 0.621 to 0.810 and for T0 and T2 from 0.603 to 0.802, indicating moderate to substantial agreement (Appendix A). Overall, vitality and wellbeing showed the highest Kappa coefficients (T0-T1=0.810; T0-T2=0.752 and T0-T1=0.767; T0-T2=0.802 respectively), while health literacy showed the lowest (T0-T1=0.621; T0-T2=0.603).

## Discussion

This study focused on the further development of the Context-specific Positive Health Questionnaire (CPHQ) (11). The objectives of this study were to evaluate the factorial, concurrent, and discriminative validity, and assess test-retest reliability. The results of the study indicate that the CPHQ2.0 is both valid and reliable. Factorial validity was confirmed, with the questionnaire clearly representing the constructs it aims to measure, showing distinct factor structures. The final questionnaire consists of 20 items distributed across 8 domains (i.e., mental relaxation, social acceptance, wellbeing, vitality, social support, financial resources, health literacy, mobility). Concurrent validity was supported by strong correlations between the CPHQ2.0 and other established questionnaire of health and well-being. The subgroup analysis revealed significant differences in scores based on factors such as gender, age, socioeconomic position (SEP), and household composition. These findings support the discriminative validity of the CPHQ2.0, indicating that the instrument can meaningfully distinguish between groups with differing health experiences. Finally, test-retest reliability demonstrated the stability of scores over time, confirming that the CPHQ2.0 is a reliable tool for measuring broad health.

The final factor structure of the CPHQ2.0 demonstrates substantial conceptual overlap with both the Capability Approach and Positive Health, and largely aligns with the eleven-factor structure of the original CPHQ (11). The CPHQ2.0 is more concise (20 versus 32 items), retaining eight factors. Three factors— political representation, environmental safety, and resilience—were excluded based on the results of the factor analysis.

Although the Capability Approach identifies political representation and perceived environmental safety as important contextual factors for health and well-being (8), these domains were not perceived as meaningful components in relation to the CPHQ2.0 by citizens, patients, and professionals. In the initial CPHQ focus group study by (12), political representation was seen as largely irrelevant to personal health: professionals viewed political matters as outside their professional remit, while citizens and patients indicated that trust in politics had little impact on their perceived health. Similarly, items relating to environmental safety led to interpretative confusion; respondents were uncertain whether “environment” referred to physical surroundings, social context, local government, or interpersonal relationships. This lack of conceptual clarity may have resulted in inconsistent responses, thereby undermining reliability of these items. These findingssuggest a potential disconnect between theoretical frameworks and how individuals subjectively interpret and experience broad health in everyday life. Future research may explore whether and how such theoretically important domains can be translated into clear, relevant, and interpretable items that resonate with people’s lived experiences.

Resilience is theoretically well-grounded in both the Capability Approach and Positive Health frameworks, regarded as a vital capacity to manage setbacks, adapt to change, and facilitate psychological recovery (30). The original CPHQ measured resilience with multiple items (11), whereas after the CPHQ focus group discussions it was reduced to one core item (“I can cope with setbacks”), supplemented by an additional item introduced by experts and professionals on setting boundaries (12). This reduction may have resulted in a loss of conceptual depth, limiting its psychometric viability as an independent factor. Resilience is a multifaceted construct that is challenging to capture comprehensively with a limited number of items. To ensure its validity in a broad health measure, resilience must be clearly defined and operationalized in a manner that is both theoretically robust and accessible to respondents (30). An alternative interpretation is that resilience may not be an inherent part of broad health itself but rather functions as a moderating factor—buffering the impact of how contextual conditions shape perceived health (31–35). Prior research has shown, for example, that resilience can reduce the negative effects of economic stress on mental well-being (34), and moderates the relationship between adverse life events and both mental and physical health (35), and are even influenced by social buffers such as social support (36). Overall, this calls for further conceptual reflection on whether—and how—resilience should be positioned within the concept of broad health: should it be elaborated as a distinct health construct, or rather be understood as a buffering mechanism that shapes how individuals experience health under adverse conditions?

A notable difference from the original CPHQ is that the social acceptance factor in CPHQ2.0 includes both positively and negatively worded items. This decision was based on focus group discussions, in which especially participants with a lower SEP identified more strongly with negatively phrased items, while professionals and experts preferred positively formulated items (11, 12). Combining both perspectives contributes to a more inclusive and realistic representation of perceived social acceptance.

The concurrent validity of the CPHQ2.0 was supported by significant correlations with various established health and well-being measures. The relatively weak correlations of the health literacy domain with other questionnaires suggest that this aspect is only marginally integrated into existing health measurement instruments. Nevertheless, within the Capability Approach, it is considered a crucial component of broad health, as it determines the extent to which individuals can meaningfully use health information in their daily lives. Findings from the focus group discussions revealed that participants found this domain difficult to comprehend, particularly when compared to more tangible domains such as vitality or financial resources (12). This aligns with the lower factor loadings and internal consistency observed for this domain. According to Pithara (2020), health literacy should be conceptualized not merely as the ability to understand information, but also to critically assess, apply, and use it in ways that align with one’s personal goals and values. Given its importance, future research should explore whether the inclusion of such broader items could enhance the conceptual clarity and reliability of this domain within the CPHQ2.0.

While it may initially seem surprising that the mental relaxation domain of the CPHQ correlates only moderately with other mental-health related questionnaires or domains, a closer examination of the individual items reveals that the CPHQ2.0 is measuring a distinct aspect of mental wellbeing. The EQ5D-5L assesses aspects of mental health in terms of anxiety and depression. The PH-17, on the other hand, is more concerned with cognitive functioning, including items like memory, concentration, and problem-solving abilities. In contrast, the CPHQ2.0 primarily addresses affective balance, reflecting individuals’ emotional state at a given moment. This distinction in focus between mental health, cognitive functioning, and affective balance helps in explaining why the mental relaxation domain of the CPHQ2.0 correlates only moderately with other domains, as it captures a different aspect of mental wellbeing. This affective aspect of wellbeing emerged as particularly important during the focus group discussions, especially among end-users (12). This emphasis aligns with the Mental Balance and Wellbeing model by Wallace and Shapiro, in which affective balance is considered the core of overall mental wellbeing (38). Other domains—cognitive, conative (motivation and intention), and attentional balance (the ability to flexibly focus and shift attention)—are seen as foundational processes that support and sustain this central affective state.

Subgroup analyses show that the CPHQ2.0 demonstrates strong discriminative validity, effectively distinguishing differences in broad health across gender, age, SEP, and household composition. These findings confirm the instrument’s sensitivity to social and demographic factors that shape health perceptions. They also underscore the importance of contextual interpretation, given the multidimensional and context-dependent nature of broad health.

The test–retest analyses demonstrated that CPHQ2.0 scores remained highly stable over a two-to four-week period, even after adjusting for significant life events between measurements. This indicates that the questionnaire is both robust and reliable for assessing broad health over short periods, while remaining sensitive enough to detect changes. The analysis further revealed that adverse life events were associated with lower levels of broad health, particularly in mental relaxation, wellbeing, and physical vitality. These findings align with earlier research by Karatzias, Jowett (2017), which showed that especially adverse life events were significantly related to decreased subjective well-being.

### Strengths and limitations

This study has several notable strengths. First, the development and validation of the CPHQ2.0 were grounded in both theoretical and empirical foundations, integrating insights from the Capability Approach and Positive Health. The sequential use of Exploratory Factor Analysis, Item Response Theory, and Confirmatory Factor Analysis allowed for a rigorous examination of the questionnaire’s factorial validity across independent data subsets, enhancing the robustness of the results. Moreover, the questionnaire development was informed by extensive input from a diverse group of stakeholders, including patients, healthcare professionals, and citizens. This participatory approach ensured that the instrument is not only theoretically sound but also practically relevant and aligned with the lived experiences of the target population.

Second, this is the first study, to our knowledge, to evaluate test-retest reliability and discriminative validity in a questionnaire designed to measure health from a broad perspective. Previous studies developing similar instruments have not yet examined measurement stability over time or systematically explored variation across demographic and health-related subgroups (10, 11, 39). Including these elements marks an important methodological advancement and takes a first step toward assessing the questionnaire’s responsiveness—its ability to detect change.

Third, the study included a test-retest reliability component over multiple time points, which, combined with linear mixed models and weighted Kappa analysis, provided a nuanced view of the temporal stability of the CPHQ2.0 while accounting for life events.

Nonetheless, limitations should also be considered. First, the cross-sectional nature of the main analyses limits causal interpretations. Although test-retest reliability was evaluated, full responsiveness to change (e.g., in response to interventions) was not assessed and should be addressed in future research.

Second, while the exclusion of domains such as political representation and perceived environmental safety was based on empirical data and end-user perspectives, it reflects a gap between theoretical models and perceived relevance in practice. Future research may explore whether and how these domains can be meaningfully translated into clear, interpretable, and reliable survey items that reflect individuals’ lived experiences of broad health.

Last, although respondents were representative of the Dutch adult population in terms of gender, socioeconomic position, and region, individuals aged 60 and older were overrepresented, which may affect the generalizability of our findings.

### Implications for practice and future research

Although the CPHQ2.0 demonstrates adequate factorial validity and internal consistency, the test–retest reliability did not exceed the commonly recommended ICC threshold of 0.90 for instruments intended for individual-level monitoring. This suggests that, in its current form, the CPHQ2.0 may not yet be suitable for closely monitoring individual health changes over time. Instead, the instrument is best suited for exploratory purposes, population-level assessments, and the evaluation of interventions, in which minor fluctuations in measurement are less critical.

Future research should further examine how resilience can best be conceptualized within the CPHQ2.0—either as an independent domain of broad health or as a moderator that influences how individuals experience health when dealing with stressors. This requires both theoretical refinement and empirical testing across diverse populations and contexts.

Furthermore, assessing the responsiveness of the CPHQ2.0 in a variety of real-world settings and across diverse population groups remains a key priority. This aligns with conclusions from a recent rapid review on the Positive Health concept (3), which highlighted the need for more evidence regarding the most suitable settings and target populations for implementation. It would also be valuable to investigate how it relates to other instruments that measure broad health concepts, such as the 17-item Positive Health questionnaire (10).

Finally, in the longer term, developing a utility-based scoring system could further enhance the instrument’s applicability in health-economic evaluations and support its use in policy and decision-making processes.

## Conclusion

The CPHQ2.0 is a valid and reliable instrument for measuring health from a broad, context-sensitive perspective. Grounded in both the Capability Approach and Positive Health, it captures key domains of perceived health based on input from diverse key-stakeholders, including citizens with a lower SEP. The questionnaire shows good factorial and concurrent validity, is sensitive to sociodemographic variation, and demonstrates temporal stability over short periods, while accounting for life events. The CPHQ2.0 is suitable for use in population-level research, health policy evaluations, and the assessment of broad health outcomes in interventions. Future research will further strengthen its utility by exploring responsiveness, refining item content, and deepening the conceptual integration of key constructs such as resilience.

## Supporting information

AppendixB_Hypotheses

AppendixA_Tables

## List of abbreviations

BRS: Brief Resilience Scale
CFA: Confirmatory Factor Analysis
CPHQ: Context-sensitive Positive Health Questionnaire
CR: Composite Reliability
EQ5D-5L: EuroQol 5 Dimensions, 5 Levels
ICC: Intraclass Correlation Coefficient
IRT: Item Response Theory
PH-17: Positive Health 17 items
PWI: Positive Wellbeing Index
SEP: Socioeconomic Position

## Declaration

### Ethics

For this study, ethical approval was obtained from the Medical Ethical Review Board of the Leiden University Medical Center (proposal number 19035).

### Consent for publication

Not applicable.

### Data availability

Materials or data used during the current study are available from the corresponding author on reasonable request.

### Author contributions

<u>Conceptualization</u>: M. Spreeuwenberg, J. Dierx, E.J. Bloemen-van Gurp, J.C. Kiefte-de Jong

<u>Formal Analysis:</u> E.M. Dubbeldeman

<u>Funding Acquisition</u>: M. Spreeuwenberg, E.J. Bloemen-van Gurp, J.C. Kiefte-de Jong

<u>Investigation</u>: M. Boelens, E.J. Bloemen-van Gurp, J.C. Kiefte-de Jong

<u>Methodology</u>: M. Spreeuwenberg, J.C. Kiefte-de Jong

<u>Project Administration</u>: M. Spreeuwenberg, J.C. Kiefte-de Jong

<u>Resources</u>: M. Boelens, E.J. Bloemen-van Gurp

<u>Supervision</u>: M. Spreeuwenberg, J. Dierx, J.C. Kiefte-de Jong

<u>Visualization:</u> E.M. Dubbeldeman

<u>Writing – Original Draft Preparation:</u> E.M. Dubbeldeman

<u>Writing – Review & Editing</u>: E.M. Dubbeldeman, M. Spreeuwenberg, J. Dierx, M. Boelens, E.J. Bloemen-van Gurp, J.C. Kiefte-de Jong

## Acknowledgments

The authors are grateful to all participants who took part in the surveys. We also thank the members of the Consortium; C. Roumen, L. Nahar-van Venrooij, M. van Vliet, M. de Kleijn, T. van Zutphen, and M. E. Akker-van Marle for their support and collaboration in this study.

## References

1. International Health Conference. Constitution of the World Health Organization. 1946. Bulletin of the World Health Organization. 2002;80(12):983–4.

2. Huber M, Knottnerus JA, Green L, Van Der Horst H, Jadad AR, Kromhout D, et al. How should we define health? Bmj. 2011;343.

3. Van Vliet M, De Kleijn M, van den Brekel-Dijkstra K, Huijts T, van Hogen-Koster S, Jung HP, et al., editors. Rapid review on the concept of positive health and its implementation in practice. Healthcare; 2024: MDPI.

4. Antonovsky A. The salutogenic model as a theory to guide health promotion. Health promotion international. 1996;11(1):11–8.

5. Mittelmark MB, Bauer GF, Vaandrager L, Pelikan JM, Sagy S, Eriksson M, et al. The handbook of salutogenesis. 2022.

6. Sen A. Capability and well-being. The quality of life. 1993;30(1):270–93.

7. Nussbaum MC. Creating capabilities: The human development approach. Harvard University Press; 2011.

8. Robeyns I, Byskov MF. The Capability Approach. In: Edward N Zalta and Uri Nodelman, editor. The Stanford Encyclopedia of Philosophy: Metaphysics Research Lab, Stanford University; 2023.

9. Prinsen CA, Terwee CB. Measuring positive health: for now, a bridge too far. Public Health. 2019;170:70–7.

10. Van Vliet M, Doornenbal BM, Boerema S, Van Den Akker-Van EM. Development and psychometric evaluation of a Positive Health measurement scale: A factor analysis study based on a Dutch population. BMJ open. 2021;11(2):e040816.

11. Doornenbal BM, van Zutphen T, Beumeler LF, Vos RC, Derks M, Haisma H, et al. Development and validation of a Context-sensitive Positive Health Questionnaire (CPHQ): A factor analysis and multivariate regression study. Journal of patient-reported outcomes. 2024;8(1):44.

12. Boelens M, Roumen C, Bloemen-van Gurp EJ, Dierx J, Nahar-van Venrooij L, van Zutphen T, et al. Stakeholder and end-user perspectives to improve the Context-sensitive Positive Health Questionnaire (CPHQ) to measure broad health. [in preparation]. 2025.

13. Flycatcher Internet Research. Flycatcher Research Agency 2024 [Available from: https://www.flycatcher.eu/nl/.

14. Von Elm E, Altman DG, Egger M, Pocock SJ, Gøtzsche PC, Vandenbroucke JP. The Strengthening the Reporting of Observational Studies in Epidemiology (STROBE) statement: guidelines for reporting observational studies. The lancet. 2007;370(9596):1453–7.

15. Comrey AL, Lee HB. A first course in factor analysis: Psychology press; 2013.

16. Worthington RL, Whittaker TA. Scale development research: A content analysis and recommendations for best practices. The counseling psychologist. 2006;34(6):806–38.

17. Herdman M, Gudex C, Lloyd A, Janssen M, Kind P, Parkin D, et al. Development and preliminary testing of the new five-level version of EQ-5D (EQ-5D-5L). Quality of life research. 2011;20:1727–36.

18. Smith BW, Dalen J, Wiggins K, Tooley E, Christopher P, Bernard J. The brief resilience scale: assessing the ability to bounce back. International journal of behavioral medicine. 2008;15:194–200.

19. Al-Janabi H, N Flynn T, Coast J. Development of a self-report measure of capability wellbeing for adults: the ICECAP-A. Quality of life research. 2012;21:167–76.

20. International Wellbeing Group. Personal Wellbeing Index. 5th edn. ed. Melbourne: Australian Centre on Quality of Life, Deakin University; 2013.

21. Schreiber JB. Issues and recommendations for exploratory factor analysis and principal component analysis. Research in Social and Administrative Pharmacy. 2021;17(5):1004–11.

22. Finch WH. Using fit statistic differences to determine the optimal number of factors to retain in an exploratory factor analysis. Educational and psychological measurement. 2020;80(2):217–41.

23. Edelen MO, Reeve BB. Applying item response theory (IRT) modeling to questionnaire development, evaluation, and refinement. Quality of life research. 2007;16:5–18.

24. Li C-H. Confirmatory factor analysis with ordinal data: Comparing robust maximum likelihood and diagonally weighted least squares. Behavior research methods. 2016;48:936–49.

25. Hooper D, Coughlan J, Mullen M. Structural equation modelling: guidelines for determining model fit. Electron J Bus Res Methods 6 (1): 53–60. 2008.

26. Shrestha N. Factor analysis as a tool for survey analysis. American journal of Applied Mathematics and statistics. 2021;9(1):4–11.

27. National Institute for Public Health and the Environment [RIVM]. Perceived Health: Employment and Income 2022 [Available from: https://www.vzinfo.nl/ervaren-gezondheid/werk-en-inkomen.

28. National Institute for Public Health and the Environment [RIVM]. Perceived Health: Causes and Consequences 2024 [Available from: https://www.vzinfo.nl/ervaren-gezondheid/oorzaken-en-gevolgen.

29. Landis J. The Measurement of Observer Agreement for Categorical Data. Biometrics. 1977.

30. Windle G. What is resilience? A review and concept analysis. Reviews in clinical gerontology. 2011;21(2):152–69.

31. Fritz J, De Graaff AM, Caisley H, Van Harmelen A-L, Wilkinson PO. A systematic review of amenable resilience factors that moderate and/or mediate the relationship between childhood adversity and mental health in young people. Frontiers in psychiatry. 2018;9:341825.

32. Zeijen ME, Brenninkmeijer V, Peeters MC, Mastenbroek NJ. The role of personal demands and personal resources in enhancing study engagement and preventing study burnout. The Spanish Journal of Psychology. 2024;27:e10.

33. Tang Y, Ma Y, Zhang J, Wang H. The relationship between negative life events and quality of life in adolescents: mediated by resilience and social support. Frontiers in public health. 2022;10:980104.

34. Essel-Gaisey F, Okyere MA, Forson R, Chiang T-F. The road to recovery: Financial resilience and mental health in post-apartheid South Africa. SSM-Population Health. 2023;23:101455.

35. Karatzias T, Jowett S, Yan E, Raeside R, Howard R. Depression and resilience mediate the relationship between traumatic life events and ill physical health: Results from a population study. Psychology, health & medicine. 2017;22(9):1021–31.

36. Ruihua L, Hassan NC, Qiuxia Z, Sha O, Jingyi D. A systematic review on the impact of social support on college students’ wellbeing and mental health. PLoS One. 2025;20(7):e0325212.

37. Pithara C. Re-thinking health literacy: using a capabilities approach perspective towards realising social justice goals. Global health promotion. 2020;27(3):150–8.

38. Wallace BA, Shapiro SL. Mental balance and well-being: building bridges between Buddhism and Western psychology. American psychologist. 2006;61(7):690.

39. Doornenbal BM, Vos RC, Van Vliet M, Kiefte-De Jong JC, van den Akker-van Marle ME. Measuring positive health: Concurrent and factorial validity based on a representative Dutch sample. Health & social care in the community. 2022;30(5):e2109–e17.

