## AppendixB_Hypotheses for "A Longitudinal Psychometric Evaluation of a Context-Sensitive Positive Health Questionnaire for Measuring Broad Health in Dutch Adults"

### Appendix X: Overview of Hypotheses for the CPHQ2.0

##### Hypothesis 1: Gender

- **Expectation**: Men will score higher (=better health) on the CPHQ2.0 compared to women.
- **Supporting Evidence**:
    1. Data from the Health Survey (CBS) shows that men consistently report better self-rated health over multiple years.
       *Source*: Self-rated health | Age and Gender | Public Health and Care (vzinfo.nl).
    2. Euroqol data indicates that men score slightly higher on the EQ-VAS (=better health) than women.
       *Source*: Population Norms for the EQ-5D - NCBI Bookshelf (nih.gov).

##### Hypothesis 2: Age

- **Expectations**:
    1. Adults of working age (18-64 years) will score higher (=better health) on the CPHQ2.0 compared to adults aged 65 and older.
    2. As participants age, scores will increase (=better health) on the CPHQ2.0 domains mental relaxation and enjoyment.
- **Supporting Evidence**:
    1. Older respondents report worse self-rated health according to the Health Survey (CBS).
       *Source*: Self-rated health | Age and Gender | Public Health and Care (vzinfo.nl).
    2. Euroqol data confirms that older adults score lower (=worse health) on the EQ-VAS compared to younger adults.
       *Source*: Population Norms for the EQ-5D - NCBI Bookshelf (nih.gov).
    3. Mental health indicators, such as calmness and happiness, improve with age.
       *Source*: Mental health issues | Public Health and Care (vzinfo.nl).

##### Hypothesis 3: Migration Background

- **Expectation**: Participants with a migration background will score lower (=worse health) on the CPHQ2.0 compared to participants with a Dutch cultural background.
- **Supporting Evidence**:
    1. A 2021 study in England involving 1,394,361 participants found that migrants scored worse on the EQ5D-5L compared to individuals without a migration background.
       *Source*: Ethnic inequalities in health-related quality of life - The Lancet Public Health.

##### Hypothesis 4: Education Level, Income, and SES

- **Expectation**: Participants with higher income, education levels, or socioeconomic status will score higher (=better health) on the CPHQ2.0 compared to those with lower income or education levels.
- **Supporting Evidence**:
    1. The Health Survey (CBS) shows that participants in higher education and income groups report better self-rated health.
       *Source*: Self-rated health | Work and Income | Public Health and Care (vzinfo.nl).
    2. Studies using ICECAP-A instruments in the Netherlands and Norway confirm these patterns.
       *Sources*:
        - The ICECAP-A instrument for capabilities - PMC (nih.gov).

**Hypothesis 5: Living Situation**

- **Expectation**: Participants who live together or are married will score higher (=better health) on the CPHQ2.0 compared to those who are single or live alone.
- **Supporting Evidence**:
  1. International studies have found differences in self-rated health/quality of life between married and single individuals.
     *Sources*:
     - Socio-economic correlates of quality of life | Health and Quality of Life Outcomes.
     - Happy, Healthy and Wedded? | PMC (nih.gov).
  2. Dutch research using the ICECAP-A instrument shows that participants in relationships score better than those without relationships.
     *Source*: The ICECAP-A instrument for capabilities - PMC (nih.gov).

**Hypothesis 6: Significant Life Events**

- **Expectations**:
  1. Participants who recently experienced life events (positive or negative) will score differently on the CPHQ2.0 compared to those who did not.
  2. There will be a cumulative effect of negative or positive life events on the CPHQ2.0 score.
- **Supporting Evidence**:
  1. A UK study (~1000 participants) found a cumulative relationship between life events and self-rated health, with negative events leading to worse outcomes and positive events leading to better outcomes.
     *Source*: The impact of life events on health and well-being - BMC Research Notes.
  2. A US study confirmed temporary changes in health scores due to negative and positive life events.
     *Source*: Exposure to negative life events - PubMed (nih.gov).
  3. Negative life events significantly impact mental well-being for both genders.
     *Source*: Gender, life events, and mental well-being - PubMed (nih.gov).

**Hypothesis 7: Chronic Conditions**

- **Expectation**: Participants with one or more chronic conditions will score lower (=worse health) on the CPHQ2.0 compared to participants without chronic conditions.
- **Supporting Evidence**:
  1. The Health Survey (CBS) shows that participants with chronic diseases report worse self-rated health than those without such conditions.
     *Source*: Self-rated health | Causes and Consequences | Public Health and Care (vzinfo.nl).
