## AppendixA_Tables for "A Longitudinal Psychometric Evaluation of a Context-Sensitive Positive Health Questionnaire for Measuring Broad Health in Dutch Adults"

### Appendix A

- **Table 1.** Socio-demographic characteristics of participants by type of dataset
- **Table 2.** Socio-demographic characteristics of participants relative to Dutch population
- **Table 3.** Parallel Analysis (including scree plot)
- **Table 4:** Model fit and variance explained for a series of Exploratory Factor Analyses
- **Table 5.** Standardized weights of the exploratory eight-factor model using oblimin rotated factors
- **Table 6a-l.** Results of the subgroup analyses: t-test and ANOVA on CPHQ scores and several characteristics of participants
- **Table 7a-i.** Results of the test-retest analyses: mixed models on CPHQ scores across time
- **Table 8.** Results of the test-retest analyses: weighted Kappa analyses on CPHQ scores

**Table 1.** Socio-demographic characteristics of participants by type of dataset

| | CFA (N=501) | EFA (N=500) | Total (N=1001) | p-value |
|---|---|---|---|---|
| **Sex** | | | | |
| Male | 244 (48.7%) | 250 (50.0%) | 494 (49.4%) | 0.919 |
| Female | 257 (51.3%) | 250 (50.0%) | 507 (50.6%) | |
| **Age** | | | | |
| 18-29 years | 80 (16.0%) | 72 (14.4%) | 152 (15.2%) | 0.913 |
| 30-44 years | 96 (19.2%) | 104 (20.8%) | 200 (20.0%) | |
| 45-59 years | 122 (24.4%) | 136 (27.2%) | 258 (25.8%) | |
| 60 years and above | 203 (40.5%) | 188 (37.6%) | 391 (39.1%) | |
| **Education level** | | | | |
| Low | 63 (12.6%) | 48 (9.6%) | 111 (11.1%) | 0.687 |
| Middle | 162 (32.3%) | 165 (33.0%) | 327 (32.7%) | |
| High | 276 (55.1%) | 287 (57.4%) | 563 (56.2%) | |
| **Income level** | | | | |
| Below modal | 115 (23.0%) | 112 (22.4%) | 227 (22.7%) | 0.999 |
| Modal | 69 (13.8%) | 71 (14.2%) | 140 (14.0%) | |
| Above modal | 313 (62.5%) | 313 (62.6%) | 626 (62.5%) | |
| Missing | 4 (0.8%) | 4 (0.8%) | 8 (0.8%) | |
| **Region of residence** | | | | |
| North | 89 (17.8%) | 96 (19.2%) | 185 (18.5%) | 0.956 |
| Middle | 288 (57.5%) | 275 (55.0%) | 563 (56.2%) | |
| South | 124 (24.8%) | 129 (25.8%) | 253 (25.3%) | |
| **SEP** | | | | |
| Low | 131 (26.1%) | 122 (24.4%) | 253 (25.3%) | 0.974 |
| High | 366 (73.1%) | 373 (74.6%) | 739 (73.8%) | |
| Unknown | 4 (0.8%) | 5 (1.0%) | 9 (0.9%) | |

*Note.* SEP = socioeconomic position.

**Table 2.** Socio-demographic characteristics of participants relative to Dutch population

| | T0 | T1 | T2 | SN 2023* |
|---|---|---|---|---|
| | **N=1001** | **N=714** | **N=543** | |
| **Sex** | | | | |
| Male | 49% | 51% | 51% | 49% |
| Female | 51% | 49% | 49% | 51% |
| **Age** | | | | |
| 18-29 years | 15% | 17% | 18% | 19% |
| 30-44 years | 20% | 22% | 21% | 23% |
| 45-59 years | 26% | 27% | 27% | 26% |
| 60 years and above | 39% | 34% | 34% | 32% |
| **Region of residence** | | | | |
| North | 19% | 19% | 19% | 19% |
| Middle | 56% | 56% | 57% | 58% |
| South | 25% | 25% | 25% | 24% |
| **SEP** | | | | |
| Low | 25% | 28% | 28% | 27% |
| High | 74% | 72% | 72% | 73% |
| Unknown | 1% | - | | - |

*Note.* *Numbers based on SN (Statistics Netherlands); SEP = socioeconomic status.

**Table 3.** Parallel analyses

| Factors | Actual | Simulated |
|---|---|---|
| 1 | 9.881641 | 0.633359 |
| 2 | 1.396466 | 0.422698 |
| 3 | 1.242953 | 0.368379 |
| 4 | 0.60676 | 0.323449 |
| 5 | 0.412056 | 0.285269 |
| 6 | 0.39626 | 0.248348 |
| 7 | 0.273896 | 0.215037 |
| 8 | 0.194532 | 0.182776 |
| 9 | 0.092237 | 0.151257 |
| 10 | 0.006139 | 0.122537 |
| 11 | -0.02995 | 0.093488 |
| 12 | -0.07773 | 0.065539 |
| 13 | -0.10872 | 0.038964 |
| 14 | -0.13986 | 0.012855 |
| 15 | -0.16474 | -0.01268 |

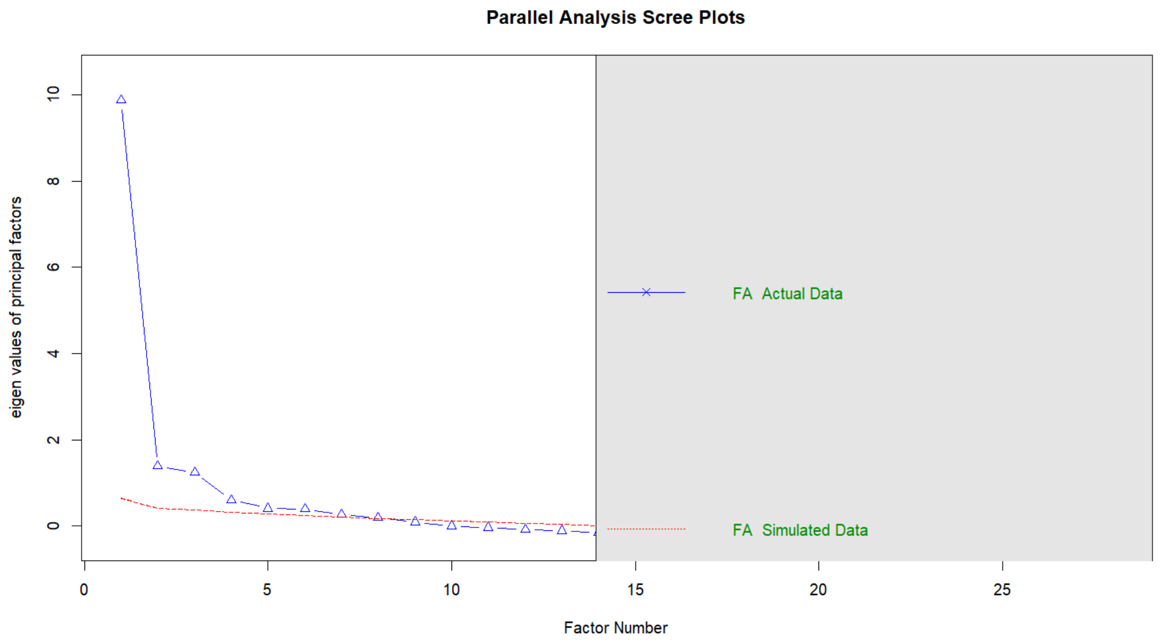

**Table 4**: Model fit and variance explained for a series of Exploratory Factor Analyses

| Number of factors | CFI | TLI | RMSEA | RMSR | Cumulative variance explained (%) |
|---|---|---|---|---|---|
| 1 | 0.66 | 0.63 | 0.12 | 0.09 | 35.30 |
| 2 | 0.76 | 0.72 | 0.10 | 0.07 | 41.14 |
| 3 | 0.83 | 0.79 | 0.09 | 0.05 | 46.40 |
| 4 | 0.87 | 0.82 | 0.08 | 0.04 | 49.25 |
| 5 | 0.91 | 0.86 | 0.07 | 0.04 | 52.38 |
| 6 | 0.93 | 0.88 | 0.07 | 0.03 | 54.72 |
| 7 | 0.95 | 0.91 | 0.06 | 0.02 | 56.74 |
| 8 | 0.97 | 0.95 | 0.05 | 0.02 | 59.16 |
| 9 | 0.98 | 0.96 | 0.04 | 0.02 | 61.04 |
| 10* | 0.99 | 0.97 | 0.04 | 0.01 | 61.86 |
| 11 | 0.97 | 0.91 | 0.06 | 0.01 | 62.99 |
| 12 | 0.99 | 0.98 | 0.03 | 0.01 | 64.22 |
| 13* | 0.99 | 0.96 | 0.04 | 0.01 | 65.30 |
| 14* | 1.00 | 0.99 | 0.02 | 0.01 | 67.31 |
| 15* | 1.00 | 0.98 | 0.03 | 0.01 | 69.22 |

*Note.* CFI = Comparative Fit Index; TLI = Tucker-Lewis Index; RMSEA = Root Mean Square Error of Approximation; SRMR = Standardized Root Mean Square Residual

**Table 5.** Standardized weights of the exploratory eight-factor model using oblimin rotated factors

| | F1 | F2 | F3 | F4 | F5 | F6 | F7 | F8 | h2 | com |
|---|---|---|---|---|---|---|---|---|---|---|
| I feel calm | **0.780** | 0.010 | -0.030 | 0.150 | 0.030 | -0.010 | 0.000 | -0.070 | 0.71 | 1.1 |
| I can relax | **0.730** | 0.030 | 0.060 | -0.060 | 0.040 | 0.050 | 0.050 | 0.090 | 0.65 | 1.1 |
| I can do what I think is important | 0.360 | 0.060 | 0.180 | -0.010 | 0.090 | 0.280 | -0.010 | 0.190 | 0.56 | 3.2 |
| I can express my boundaries | 0.350 | 0.180 | -0.080 | 0.120 | -0.020 | -0.120 | 0.330 | -0.150 | 0.36 | 3.5 |
| I can cope with setbacks | 0.340 | 0.100 | 0.100 | 0.060 | -0.050 | -0.030 | 0.320 | -0.080 | 0.38 | 2.6 |
| I feel accepted | 0.020 | **0.730** | 0.090 | -0.050 | -0.040 | 0.120 | 0.050 | 0.070 | 0.71 | 1.1 |
| I feel that I belong | 0.000 | **0.670** | 0.030 | 0.070 | 0.250 | -0.040 | 0.010 | -0.020 | 0.74 | 1.3 |
| I feel excluded* | 0.070 | **0.580** | 0.070 | 0.060 | 0.100 | 0.040 | -0.030 | -0.050 | 0.54 | 1.2 |
| I feel happy | 0.170 | 0.080 | **0.600** | 0.120 | 0.080 | 0.050 | -0.030 | -0.020 | 0.72 | 1.3 |
| I feel my life is meaningful | -0.040 | 0.170 | **0.590** | 0.160 | 0.060 | -0.020 | 0.100 | -0.070 | 0.65 | 1.4 |
| I feel grateful for my life | -0.070 | 0.210 | **0.510** | 0.080 | 0.060 | 0.030 | 0.070 | 0.000 | 0.50 | 1.5 |
| I can enjoy | 0.360 | 0.050 | **0.450** | -0.160 | 0.050 | 0.040 | 0.060 | 0.250 | 0.65 | 3.0 |
| I have goals and ideals I want to achieve | -0.090 | -0.240 | 0.280 | 0.210 | 0.240 | 0.110 | 0.260 | -0.100 | 0.37 | 5.7 |
| I feel fit | 0.060 | 0.040 | 0.010 | **0.800** | -0.020 | 0.070 | -0.020 | 0.120 | 0.81 | 1.1 |
| I feel healthy | 0.050 | -0.010 | 0.100 | **0.760** | 0.000 | 0.060 | 0.040 | 0.050 | 0.77 | 1.1 |
| I feel good about myself | 0.380 | 0.040 | 0.270 | 0.390 | 0.090 | 0.010 | -0.050 | -0.050 | 0.74 | 3.0 |
| I have people who support me | 0.010 | 0.000 | -0.010 | -0.030 | **0.900** | 0.020 | 0.020 | 0.000 | 0.83 | 1.0 |
| I feel that people support me | 0.030 | 0.300 | -0.030 | -0.020 | **0.560** | 0.050 | 0.100 | -0.020 | 0.69 | 1.7 |
| I have people with I can do fun things with | 0.040 | 0.060 | 0.120 | 0.000 | **0.550** | 0.060 | -0.100 | 0.140 | 0.49 | 1.4 |
| I have money to do the things that I find important | 0.000 | 0.000 | -0.020 | 0.000 | 0.000 | **1.020** | -0.030 | -0.030 | 0.99 | 1.0 |
| I have money to live a healthy life | 0.000 | 0.030 | -0.010 | 0.050 | 0.010 | **0.760** | 0.070 | 0.010 | 0.66 | 1.0 |
| I feel safe where I live | -0.060 | 0.220 | 0.130 | -0.030 | -0.090 | 0.280 | 0.190 | 0.090 | 0.29 | 3.8 |
| I can understand information about my health | 0.040 | 0.060 | -0.040 | -0.120 | 0.000 | 0.120 | **0.570** | 0.080 | 0.41 | 1.3 |
| I can find help for my health | 0.020 | -0.040 | 0.080 | 0.010 | 0.150 | 0.090 | **0.550** | 0.090 | 0.52 | 1.3 |
| I can move | 0.010 | 0.080 | -0.090 | 0.330 | 0.050 | 0.030 | 0.050 | **0.610** | 0.67 | 1.7 |
| I can be active in my environment | -0.010 | -0.030 | 0.100 | 0.220 | 0.110 | 0.030 | 0.260 | **0.430** | 0.58 | 2.6 |
| I can work/volunteer or provide informal care | -0.080 | 0.010 | 0.080 | 0.230 | 0.090 | 0.000 | 0.120 | 0.320 | 0.30 | 2.6 |
| I have confidence in society | 0.050 | 0.190 | 0.120 | 0.140 | 0.040 | 0.110 | 0.190 | -0.260 | 0.30 | 4.6 |

*Note.* * Negative formulation; F1 = Mental Relaxation; F2 = Social Acceptance; F3 = Enjoyment; F4 = Fitness; F5 = Social Support; F6 = Financial Resources; F7 = Health Literacy; F8 = Autonomy.

**Table 6a.** Results of subgroup analysis: independent samples t-test on CPHQ scores and gender

| | Male (N=494) | Female (N=507) | Total (N=1001) | p-value |
|---|---|---|---|---|
| **CPHQ mental relaxation** | | | | <0.001 |
| Mean (SD) | 73.1 (16.3) | 68.2 (16.7) | 70.6 (16.7) | |
| **CPHQ social acceptance** | | | | <0.001 |
| Mean (SD) | 78.3 (13.5) | 74.5 (16.2) | 76.4 (15.0) | |
| **CPHQ enjoyment** | | | | 0.146 |
| Mean (SD) | 75.5 (14.8) | 74.1 (15.3) | 74.8 (15.0) | |
| **CPHQ fitness** | | | | 0.092 |
| Mean (SD) | 67.8 (19.1) | 65.8 (20.0) | 66.8 (19.6) | |
| **CPHQ social support** | | | | 0.107 |
| Mean (SD) | 75.4 (13.3) | 76.8 (14.8) | 76.1 (14.1) | |
| **CPHQ financial resources** | | | | 0.483 |
| Mean (SD) | 80.3 (16.4) | 79.6 (17.6) | 80.0 (17.0) | |
| **CPHQ health literacy** | | | | 0.909 |
| Mean (SD) | 81.4 (11.8) | 81.3 (13.0) | 81.3 (12.4) | |
| **CPHQ autonomy** | | | | 0.710 |
| Mean (SD) | 77.0 (14.3) | 77.3 (14.9) | 77.2 (14.6) | |
| **CPHQ total score** | | | | 0.045 |
| Mean (SD) | 76.1 (10.5) | 74.7 (11.5) | 75.4 (11.0) | |

**Table 6b.** Results of subgroup analysis: independent samples t-test on CPHQ scores and age

| | 18-64 (N=728) | 65 and above (N=273) | Total (N=1001) | p-value |
|---|---|---|---|---|
| **CPHQ mental relaxation** | | | | <0.001 |
| Mean (SD) | 69.4 (17.4) | 73.9 (14.0) | 70.6 (16.7) | |
| **CPHQ social acceptance** | | | | 0.014 |
| Mean (SD) | 75.7 (15.7) | 78.1 (13.1) | 76.4 (15.0) | |
| **CPHQ enjoyment** | | | | 0.714 |
| Mean (SD) | 74.7 (15.6) | 75.0 (13.3) | 74.8 (15.0) | |
| **CPHQ fitness** | | | | 0.033 |
| Mean (SD) | 67.6 (19.7) | 64.7 (19.1) | 66.8 (19.6) | |
| **CPHQ social support** | | | | <0.001 |
| Mean (SD) | 77.0 (14.3) | 73.7 (13.3) | 76.1 (14.1) | |
| **CPHQ financial resources** | | | | 0.016 |
| Mean (SD) | 80.8 (16.9) | 77.8 (17.1) | 80.0 (17.0) | |
| **CPHQ health literacy** | | | | 0.003 |
| Mean (SD) | 82.0 (12.9) | 79.5 (11.0) | 81.3 (12.4) | |
| **CPHQ autonomy** | | | | <0.001 |
| Mean (SD) | 78.5 (14.2) | 73.6 (15.1) | 77.2 (14.6) | |
| **CPHQ total score** | | | | 0.125 |
| Mean (SD) | 75.7 (11.3) | 74.6 (10.2) | 75.4 (11.0) | |

**Table 6c.** Results of subgroup analysis independent samples t-test on CPHQ scores and migration background

| | Both parents born in the Netherlands (N=962) | Both parents born outside the Netherlands (N=36) | Total (N=998) | p-value |
|---|---|---|---|---|
| **CPHQ mental relaxation** | | | | 0.069 |
| Mean (SD) | 71.0 (16.5) | 67.3 (18.4) | 70.7 (16.7) | |
| **CPHQ social acceptance** | | | | 0.069 |
| Mean (SD) | 76.7 (14.8) | 73.4 (16.3) | 76.4 (15.0) | |
| **CPHQ enjoyment** | | | | 0.153 |
| Mean (SD) | 75.1 (14.8) | 72.5 (16.4) | 74.8 (15.0) | |
| **CPHQ fitness** | | | | 0.112 |
| Mean (SD) | 67.2 (19.5) | 63.6 (19.9) | 66.8 (19.5) | |
| **CPHQ social support** | | | | 0.104 |
| Mean (SD) | 76.4 (14.2) | 74.0 (13.0) | 76.2 (14.1) | |
| **CPHQ financial resources** | | | | 0.010 |
| Mean (SD) | 80.4 (16.9) | 75.4 (17.1) | 80.0 (17.0) | |
| **CPHQ health literacy** | | | | 0.011 |
| Mean (SD) | 81.7 (12.2) | 77.7 (14.3) | 81.4 (12.4) | |
| **CPHQ autonomy** | | | | 0.040 |
| Mean (SD) | 77.5 (14.6) | 74.2 (14.6) | 77.2 (14.6) | |
| **CPHQ total score** | | | | 0.007 |
| Mean (SD) | 75.7 (10.9) | 72.2 (11.5) | 75.4 (11.0) | |

**Table 6d.** Results of subgroup analysis: ANOVA on CPHQ scores and educational level

| | Low (N=111) | Middle (N=327) | High (N=563) | Total (N=1001) | p-value |
|---|---|---|---|---|---|
| **CPHQ mental relaxation** | | | | | 0.173 |
| Mean (SD) | 68.1 (16.2) | 71.6 (17.1) | 70.6 (16.5) | 2.17 (0.667) | |
| **CPHQ social acceptance** | | | | | <0.001 |
| Mean (SD) | 71.5 (15.5) | 76.1 (15.3) | 77.4 (14.6) | 1.95 (0.602) | |
| **CPHQ enjoyment** | | | | | <0.001 |
| Mean (SD) | 69.3 (16.0) | 74.7 (16.1) | 75.9 (13.9) | 2.01 (0.601) | |
| **CPHQ fitness** | | | | | <0.001 |
| Mean (SD) | 55.5 (20.1) | 65.1 (19.5) | 70.0 (18.6) | 2.33 (0.783) | |
| **CPHQ social support** | | | | | <0.001 |
| Mean (SD) | 67.9 (15.2) | 75.4 (14.1) | 78.2 (13.3) | 1.96 (0.565) | |
| **CPHQ financial resources** | | | | | <0.001 |
| Mean (SD) | 64.8 (18.2) | 76.5 (17.5) | 85.0 (13.9) | 1.80 (0.680) | |
| **CPHQ health literacy** | | | | | <0.001 |
| Mean (SD) | 73.9 (10.0) | 79.1 (12.2) | 84.1 (12.1) | 1.75 (0.497) | |
| **CPHQ autonomy** | | | | | <0.001 |
| Mean (SD) | 68.1 (14.4) | 76.1 (14.8) | 79.6 (13.7) | 1.91 (0.584) | |
| **CPHQ total score** | | | | | <0.001 |
| Mean (SD) | 67.4 (10.7) | 74.3 (11.3) | 77.6 (10.1) | 1.98 (0.441) | |

**Table 6e.** Results of subgroup analysis: ANOVA on CPHQ scores and income

| | Below modal (N=227) | Modal (N=140) | Above modal (N=626) | Total (N=993) | p-value |
|---|---|---|---|---|---|
| **CPHQ mental relaxation** | | | | | 0.097 |
| Mean (SD) | 68.6 (16.7) | 71.4 (16.4) | 71.2 (16.8) | 70.7 (16.7) | |
| **CPHQ social acceptance** | | | | | <0.001 |
| Mean (SD) | 71.4 (15.9) | 76.5 (13.3) | 78.0 (14.8) | 76.3 (15.1) | |
| **CPHQ enjoyment** | | | | | <0.001 |
| Mean (SD) | 69.9 (17.0) | 76.5 (13.6) | 76.2 (14.3) | 74.8 (15.1) | |
| **CPHQ fitness** | | | | | <0.001 |
| Mean (SD) | 58.3 (21.4) | 67.9 (19.2) | 69.7 (17.9) | 66.9 (19.5) | |
| **CPHQ social support** | | | | | <0.001 |
| Mean (SD) | 70.5 (15.7) | 75.2 (12.1) | 78.4 (13.4) | 76.1 (14.1) | |
| **CPHQ financial resources** | | | | | <0.001 |
| Mean (SD) | 66.5 (19.2) | 79.2 (15.0) | 85.0 (13.6) | 79.9 (17.0) | |
| **CPHQ health literacy** | | | | | <0.001 |
| Mean (SD) | 76.5 (11.9) | 81.3 (13.4) | 83.1 (12.0) | 81.3 (12.5) | |
| **CPHQ autonomy** | | | | | <0.001 |
| Mean (SD) | 70.5 (16.0) | 78.3 (14.4) | 79.4 (13.3) | 77.2 (14.6) | |
| **CPHQ total score** | | | | | <0.001 |
| Mean (SD) | 69.0 (12.0) | 75.8 (10.3) | 77.6 (9.93) | 75.4 (11.1) | |

**Table 6f.** Results of subgroup analysis: independent samples t-test on CPHQ scores and socioeconomic status

| | **Low (N=253)** | **High (N=739)** | **Total (N=992)** | **p-value** |
|---|---|---|---|---|
| **CPHQ mental relaxation** | | | | 0.056 |
| Mean (SD) | 68.9 (16.8) | 71.3 (16.7) | 70.7 (16.7) | |
| **CPHQ social acceptance** | | | | <0.001 |
| Mean (SD) | 72.4 (15.7) | 77.7 (14.6) | 76.3 (15.1) | |
| **CPHQ enjoyment** | | | | <0.001 |
| Mean (SD) | 70.7 (16.8) | 76.2 (14.2) | 74.8 (15.1) | |
| **CPHQ fitness** | | | | <0.001 |
| Mean (SD) | 58.3 (21.0) | 69.8 (18.1) | 66.9 (19.5) | |
| **CPHQ social support** | | | | <0.001 |
| Mean (SD) | 70.9 (15.3) | 77.9 (13.3) | 76.1 (14.1) | |
| **CPHQ financial resources** | | | | <0.001 |
| Mean (SD) | 67.3 (18.8) | 84.3 (14.0) | 79.9 (17.0) | |
| **CPHQ health literacy** | | | | <0.001 |
| Mean (SD) | 76.3 (11.7) | 83.1 (12.3) | 81.4 (12.5) | |
| **CPHQ autonomy** | | | | <0.001 |
| Mean (SD) | 70.8 (15.8) | 79.4 (13.5) | 77.2 (14.6) | |
| **CPHQ total score** | | | | <0.001 |
| Mean (SD) | 69.5 (11.8) | 77.4 (10.0) | 75.4 (11.1) | |

**Table 6g.** Results of subgroup analysis: independent samples t-test on CPHQ scores and household composition

| | Without partner (N=276) | With partner (N=725) | Total (N=1001) | p-value |
|---|---|---|---|---|
| **CPHQ mental relaxation** | | | | 0.160 |
| Mean (SD) | 69.4 (17.9) | 71.1 (16.2) | 70.6 (16.7) | |
| **CPHQ social acceptance** | | | | <0.001 |
| Mean (SD) | 72.5 (16.1) | 77.8 (14.4) | 76.4 (15.0) | |
| **CPHQ enjoyment** | | | | <0.001 |
| Mean (SD) | 70.9 (16.4) | 76.3 (14.2) | 74.8 (15.0) | |
| **CPHQ social support** | | | | <0.001 |
| Mean (SD) | 73.4 (15.8) | 77.2 (13.3) | 76.1 (14.1) | |
| **CPHQ financial resources** | | | | <0.001 |
| Mean (SD) | 76.0 (19.7) | 81.5 (15.6) | 80.0 (17.0) | |
| **CPHQ health literacy** | | | | 0.172 |
| Mean (SD) | 80.4 (13.0) | 81.7 (12.2) | 81.3 (12.4) | |
| **CPHQ autonomy** | | | | 0.152 |
| Mean (SD) | 76.0 (15.9) | 77.6 (14.1) | 77.2 (14.6) | |
| **CPHQ total score** | | | | <0.001 |
| Mean (SD) | 72.9 (12.4) | 76.3 (10.3) | 75.4 (11.0) | |

**Table 6h.** Results of subgroup analysis: independent samples t-test on CPHQ scores and adverse life events

| | **No (N=571)** | **Yes (N=430)** | **Total (N=1001)** | **p-value** |
|---|---|---|---|---|
| **CPHQ mental relaxation** | | | | <0.001 |
| Mean (SD) | 73.2 (15.3) | 67.2 (17.8) | 70.6 (16.7) | |
| **CPHQ social acceptance** | | | | 0.047 |
| Mean (SD) | 77.2 (14.4) | 75.3 (15.8) | 76.4 (15.0) | |
| **CPHQ enjoyment** | | | | 0.007 |
| Mean (SD) | 75.9 (14.5) | 73.3 (15.5) | 74.8 (15.0) | |
| **CPHQ social support** | | | | 0.897 |
| Mean (SD) | 76.1 (14.0) | 76.2 (14.3) | 76.1 (14.1) | |
| **CPHQ financial resources** | | | | 0.692 |
| Mean (SD) | 80.1 (16.5) | 79.7 (17.7) | 80.0 (17.0) | |
| **CPHQ health literacy** | | | | 0.620 |
| Mean (SD) | 81.5 (12.1) | 81.1 (12.9) | 81.3 (12.4) | |
| **CPHQ autonomy** | | | | 0.004 |
| Mean (SD) | 78.3 (13.8) | 75.6 (15.5) | 77.2 (14.6) | |
| **CPHQ total score** | | | | <0.001 |
| Mean (SD) | 76.5 (10.4) | 73.9 (11.7) | 75.4 (11.0) | |

**Table 6i.** Results of subgroup analysis: ANOVA on CPHQ scores and number of adverse life events

| | No life events (N=571) | One life event (N=300) | 2 or more life events (N=130) | Total (N=1001) | p-value |
|---|---|---|---|---|---|
| **CPHQ mental relaxation** | | | | | <0.001 |
| Mean (SD) | 73.2 (15.3) | 68.4 (17.2) | 64.3 (18.7) | 70.6 (16.7) | |
| **CPHQ social acceptance** | | | | | 0.026 |
| Mean (SD) | 77.2 (14.4) | 76.1 (15.1) | 73.3 (17.3) | 76.4 (15.0) | |
| **CPHQ enjoyment** | | | | | <0.001 |
| Mean (SD) | 75.9 (14.5) | 74.5 (14.8) | 70.4 (16.8) | 74.8 (15.0) | |
| **CPHQ fitness** | | | | | <0.001 |
| Mean (SD) | 69.7 (18.3) | 64.2 (20.6) | 59.8 (20.0) | 66.8 (19.6) | |
| **CPHQ social support** | | | | | 0.590 |
| Mean (SD) | 76.1 (14.0) | 76.6 (13.6) | 75.1 (15.8) | 76.1 (14.1) | |
| **CPHQ financial resources** | | | | | 0.726 |
| Mean (SD) | 80.1 (16.5) | 80.1 (17.2) | 78.8 (18.9) | 80.0 (17.0) | |
| **CPHQ health literacy** | | | | | 0.423 |
| Mean (SD) | 81.5 (12.1) | 81.6 (12.2) | 80.0 (14.5) | 81.3 (12.4) | |
| **CPHQ autonomy** | | | | | 0.003 |
| Mean (SD) | 78.3 (13.8) | 76.4 (14.9) | 73.8 (16.7) | 77.2 (14.6) | |
| **CPHQ total score** | | | | | <0.001 |
| Mean (SD) | 76.5 (10.4) | 74.7 (11.2) | 72.0 (12.6) | 75.4 (11.0) | |

**Table 6j.** Results of subgroup analysis: independent samples t-test on CPHQ scores and positive life events

| | No (N=754) | Yes (N=247) | Total (N=1001) | p-value |
|---|---|---|---|---|
| **CPHQ mental relaxation** | | | | 0.121 |
| Mean (SD) | 71.1 (16.8) | 69.2 (16.3) | 70.6 (16.7) | |
| **CPHQ social acceptance** | | | | 0.433 |
| Mean (SD) | 76.6 (15.1) | 75.7 (14.9) | 76.4 (15.0) | |
| **CPHQ enjoyment** | | | | 0.289 |
| Mean (SD) | 74.5 (14.8) | 75.7 (15.6) | 74.8 (15.0) | |
| **CPHQ fitness** | | | | 0.078 |
| Mean (SD) | 66.2 (19.6) | 68.7 (19.3) | 66.8 (19.6) | |
| **CPHQ social support** | | | | 0.017 |
| Mean (SD) | 75.5 (14.0) | 78.0 (14.4) | 76.1 (14.1) | |
| **CPHQ financial resources** | | | | 0.288 |
| Mean (SD) | 79.7 (17.7) | 80.9 (14.8) | 80.0 (17.0) | |
| **CPHQ health literacy** | | | | 0.044 |
| Mean (SD) | 80.9 (12.5) | 82.7 (12.1) | 81.3 (12.4) | |
| **CPHQ autonomy** | | | | 0.038 |
| Mean (SD) | 76.6 (14.8) | 78.8 (13.9) | 77.2 (14.6) | |
| **CPHQ total score** | | | | 0.174 |
| Mean (SD) | 75.1 (11.1) | 76.2 (10.7) | 75.4 (11.0) | |

**Table 6k.** Results of subgroup analysis: ANVOA on CPHQ scores and number of positive life events

| | No life events (N=754) | One life event (N=91) | 2 or more life events (N=156) | Total (N=1001) | p-value |
|---|---|---|---|---|---|
| **CPHQ mental relaxation** | | | | | 0.244 |
| Mean (SD) | 71.1 (16.8) | 70.2 (16.8) | 68.7 (16.0) | 70.6 (16.7) | |
| **CPHQ social acceptance** | | | | | 0.557 |
| Mean (SD) | 76.6 (15.1) | 76.6 (12.6) | 75.2 (16.1) | 76.4 (15.0) | |
| **CPHQ enjoyment** | | | | | 0.546 |
| Mean (SD) | 74.5 (14.8) | 75.9 (15.3) | 75.6 (15.8) | 74.8 (15.0) | |
| **CPHQ fitness** | | | | | 0.131 |
| Mean (SD) | 66.2 (19.6) | 67.0 (19.2) | 69.6 (19.4) | 66.8 (19.6) | |
| **CPHQ social support** | | | | | 0.040 |
| Mean (SD) | 75.5 (14.0) | 77.1 (14.4) | 78.5 (14.5) | 76.1 (14.1) | |
| **CPHQ financial resources** | | | | | 0.619 |
| Mean (SD) | 79.7 (17.7) | 81.0 (15.5) | 80.8 (14.4) | 80.0 (17.0) | |
| **CPHQ health literacy** | | | | | 0.019 |
| Mean (SD) | 80.9 (12.5) | 80.6 (11.7) | 83.9 (12.2) | 81.3 (12.4) | |
| **CPHQ autonomy** | | | | | 0.100 |
| Mean (SD) | 76.6 (14.8) | 77.9 (15.9) | 79.3 (12.5) | 77.2 (14.6) | |
| **CPHQ total score** | | | | | 0.372 |
| Mean (SD) | 75.1 (11.1) | 75.8 (11.1) | 76.4 (10.6) | 75.4 (11.0) | |

**Table 6l.** Results of subgroup analysis: independent samples t-test on CPHQ scores and the presence of non-communicable diseases

| | **No (N=482)** | **Yes (N=519)** | **Total (N=1001)** | **p-value** |
|---|---|---|---|---|
| **CPHQ mental relaxation** | | | | <0.001 |
| Mean (SD) | 2.09 (0.639) | 2.25 (0.685) | 2.17 (0.667) | |
| **CPHQ social acceptance** | | | | <0.001 |
| Mean (SD) | 1.88 (0.585) | 2.01 (0.610) | 1.95 (0.602) | |
| **CPHQ enjoyment** | | | | <0.001 |
| Mean (SD) | 1.91 (0.553) | 2.10 (0.629) | 2.01 (0.601) | |
| **CPHQ fitness** | | | | <0.001 |
| Mean (SD) | 2.02 (0.574) | 2.61 (0.842) | 2.33 (0.783) | |
| **CPHQ social support** | | | | 0.00177 |
| Mean (SD) | 1.90 (0.551) | 2.01 (0.573) | 1.96 (0.565) | |
| **CPHQ financial resources** | | | | <0.001 |
| Mean (SD) | 1.70 (0.565) | 1.89 (0.762) | 1.80 (0.680) | |
| **CPHQ health literacy** | | | | <0.001 |
| Mean (SD) | 1.69 (0.465) | 1.80 (0.520) | 1.75 (0.497) | |
| **CPHQ autonomy** | | | | <0.001 |
| Mean (SD) | 1.75 (0.502) | 2.06 (0.615) | 1.91 (0.584) | |
| **CPHQ total score** | | | | <0.001 |
| Mean (SD) | 1.87 (0.375) | 2.09 (0.471) | 1.98 (0.441) | |

**Table 7a.** Results of the test-retest analysis: mixed models for CPHQ mental relaxation

| Effect | Estimate | se | t | p-value | 95% CI |
|---|---|---|---|---|---|
| **Intercept** | 74.444 | 1.061 | 70.141 | < .001 | [72.359 – 76.528] |
| **Time [ref = T0]** | | | | | |
| T1 | -0.184 | 0.515 | -0.357 | 0.721 | [-1.197 – 0.828] |
| T2 | -0.391 | 0.485 | -0.806 | 0.420 | [-1.344 – 0.561] |
| **Adverse events [ref = No]** | | | | | |
| Event at T0 | -6.007 | 1.417 | -4.239 | < .001 | [-8.790 – -3.223] |
| Event at T1 | -3.693 | 1.895 | -1.949 | 0.052 | [-7.416 – 0.030] |
| Event at T2 | -2.762 | 1.840 | -1.501 | 0.134 | [-6.376 – 0.852] |
| **Positive events [ref = No]** | | | | | |
| Event at T0 | -0.496 | 1.538 | -0.322 | 0.747 | [-3.517 – 2.526] |
| Event at T1 | 0.455 | 3.449 | 0.132 | 0.895 | [-6.320 – 7.231] |
| Event at T2 | 1.049 | 4.932 | 0.213 | 0.832 | [-8.639 – 10.737] |
| **Covariance parameters** | **Estimate** | **se** | **Wald Z** | **p-value** | **95% CI** |
| Between variance | 221.603 | 16.072 | 13.788 | <.001 | [192.239 – 255.452] |
| Within variance | 73.308 | 5.520 | 13.281 | <.001 | [63.250 – 84.966] |
| Intraclass correlation | 0.751 | - | - | - | - |

**Table 7b.** Results of the test-retest analysis: mixed models for CPHQ social acceptance

| Effect | Estimate | se | t | p-value | 95% CI |
|---|---|---|---|---|---|
| **Intercept** | 77.357 | 0.924 | 83.752 | < .001 | [75.543 – 79.171] |
| **Time [ref = T0]** | | | | | |
| T1 | -0.844 | 0.416 | -2.030 | 0.043 | [-1.661 – -0.027] |
| T2 | -0.061 | 0.393 | -0.156 | 0.876 | [-0.832 – 0.709] |
| **Adverse events [ref = No]** | | | | | |
| Event at T0 | -2.416 | 1.240 | -1.949 | 0.052 | [-4.852 – 0.019] |
| Event at T1 | -2.259 | 1.658 | -1.363 | 0.174 | [-5.517 – 0.998] |
| Event at T2 | -1.618 | 1.610 | -1.005 | 0.315 | [-4.780 – 1.545] |
| **Positive events [ref = No]** | | | | | |
| Event at T0 | -0.739 | 1.346 | -0.549 | 0.583 | [-3.382 – 1.905] |
| Event at T1 | 0.983 | 3.018 | 0.326 | 0.745 | [-4.945 – 6.912] |
| Event at T2 | 0.214 | 4.315 | 0.050 | 0.960 | [-8.262 – 8.691] |
| **Covariance parameters** | **Estimate** | **se** | **Wald Z** | **p-value** | **95% CI** |
| Between variance | 173.116 | 11.992 | 14.436 | < .001 | [151.137 – 198.291] |
| Within variance | 47.643 | 3.430 | 13.890 | < .001 | [41.373 – 54.863] |
| Intraclass correlation | 0.784 | - | - | - | - |

**Table 7c.** Results of the test-retest analysis: mixed models for CPHQ enjoyment

| Effect | Estimate | se | t | p-value | 95% CI |
|---|---|---|---|---|---|
| **Intercept** | 76.070 | 0.980 | 77.627 | < .001 | [74.145 – 77.994] |
| **Time [ref = T0]** | | | | | |
| T1 | -0.127 | 0.369 | -0.343 | 0.732 | [-0.852 – 0.599] |
| T2 | -0.219 | 0.376 | -0.582 | 0.561 | [-0.957 – 0.519] |
| **Adverse events [ref = No]** | | | | | |
| Event at T0 | -3.066 | 1.325 | -2.314 | 0.021 | [-5.670 – -0.463] |
| Event at T1 | -0.972 | 1.773 | -0.548 | 0.584 | [-4.454 – 2.510] |
| Event at T2 | -3.316 | 1.721 | -1.927 | 0.055 | [-6.696 – 0.065] |
| **Positive events [ref = No]** | | | | | |
| Event at T0 | 0.679 | 1.439 | 0.472 | 0.637 | [-2.147 – 3.506] |
| Event at T1 | 1.941 | 3.226 | 0.602 | 0.548 | [-4.396 – 8.278] |
| Event at T2 | 5.090 | 4.613 | 1.104 | 0.270 | [-3.971 – 14.152] |
| **Covariance parameters** | **Estimate** | **se** | **Wald Z** | **p-value** | **95% CI** |
| Between variance | 207.242 | 13.461 | 15.396 | < .001 | [182.470 – 235.377] |
| Within variance | 37.077 | 2.122 | 17.472 | < .001 | [33.143 – 41.478] |
| Intraclass correlation | 0.848 | - | - | - | - |

**Table 7d.** Results of the test-retest analysis: mixed models for CPHQ fitness

| Effect | Estimate | se | t | p-value | 95% CI |
|---|---|---|---|---|---|
| **Intercept** | 69.979 | 1.194 | 58.632 | < .001 | [67.635 – 72.323] |
| **Time [ref = T0]** | | | | | |
| T1 | -0.161 | 0.538 | -0.300 | 0.765 | [-1.218 – 0.896] |
| T2 | -0.829 | 0.491 | -1.687 | 0.092 | [-1.793 – 0.135] |
| **Adverse events [ref = No]** | | | | | |
| Event at T0 | -5.861 | 1.604 | -3.655 | < .001 | [-9.012 – -2.711] |
| Event at T1 | -3.846 | 2.145 | -1.793 | 0.074 | [-8.061 – 0.368] |
| Event at T2 | -5.391 | 2.083 | -2.588 | 0.010 | [-9.483 – -1.300] |
| **Positive events [ref = No]** | | | | | |
| Event at T0 | 1.886 | 1.741 | 1.083 | 0.279 | [-1.534 – 5.307] |
| Event at T1 | 2.132 | 3.905 | 0.546 | 0.585 | [-5.538 – 9.802] |
| Event at T2 | 7.256 | 5.583 | 1.300 | 0.194 | [-3.710 – 18.222] |
| **Covariance parameters** | **Estimate** | **se** | **Wald Z** | **p-value** | **95% CI** |
| Between variance | 285.684 | 20.398 | 14.009 | < .001 | [248.448 – 328.667] |
| Within variance | 81.857 | 6.808 | 12.024 | < .001 | [69.544 – 96.350] |
| Intraclass correlation | 0.777 | - | - | - | - |

**Table 7e.** Results of the test-retest analysis: mixed models for CPHQ social support

| Effect | Estimate | se | t | p-value | 95% CI |
|---|---|---|---|---|---|
| **Intercept** | 75.343 | 0.874 | 86.253 | < .001 | [73.628 – 77.059] |
| **Time [ref = T0]** | | | | | |
| T1 | -0.476 | 0.419 | -1.136 | 0.257 | [-1.299 – 0.347] |
| T2 | -0.338 | 0.365 | -0.925 | 0.355 | [-1.054 – 0.379] |
| **Adverse events [ref = No]** | | | | | |
| Event at T0 | 0.012 | 1.171 | 0.010 | 0.992 | [-2.288 – 2.312] |
| Event at T1 | 0.002 | 1.566 | 0.001 | 0.999 | [-3.075 – 3.079] |
| Event at T2 | -2.814 | 1.521 | -1.850 | 0.065 | [-5.801 – 0.173] |
| **Positive events [ref = No]** | | | | | |
| Event at T0 | 2.363 | 1.271 | 1.859 | 0.064 | [-0.135 – 4.860] |
| Event at T1 | 2.933 | 2.851 | 1.029 | 0.304 | [-2.667 – 8.532] |
| Event at T2 | -0.477 | 4.076 | -0.117 | 0.907 | [-8.484 – 7.529] |
| **Covariance parameters** | **Estimate** | **se** | **Wald Z** | **p-value** | **95% CI** |
| Between variance | 145.020 | 11.468 | 12.646 | < .001 | [124.198 – 169.332] |
| Within variance | 52.958 | 5.443 | 9.729 | < .001 | [43.296 – 64.778] |
| Intraclass correlation | 0.733 | - | - | - | - |

**Table 7f.** Results of the test-retest analysis: mixed models for CPHQ financial resources

| Effect | Estimate | se | t | p-value | 95% CI |
|---|---|---|---|---|---|
| **Intercept** | 79.318 | 1.061 | 74.748 | < .001 | [77.234 – 81.402] |
| **Time [ref = T0]** | | | | | |
| T1 | -0.622 | 0.495 | -1.256 | 0.210 | [-1.594 – 0.351] |
| T2 | -0.737 | 0.448 | -1.644 | 0.101 | [-1.616 – 0.143] |
| **Adverse events [ref = No]** | 0b | 0 | . | . | . |
| Event at T0 | | | | | |
| Event at T1 | -1.016 | 1.423 | -0.714 | 0.476 | [-3.811 – 1.780] |
| Event at T2 | 0.298 | 1.904 | 0.156 | 0.876 | [-3.442 – 4.037] |
| **Positive events [ref = No]** | -2.281 | 1.848 | -1.234 | 0.218 | [-5.911 – 1.350] |
| Event at T0 | | | | | |
| Event at T1 | 1.715 | 1.545 | 1.110 | 0.268 | [-1.321 – 4.750] |
| Event at T2 | 0.346 | 3.465 | 0.100 | 0.920 | [-6.460 – 7.152] |
| **Covariance parameters** | **Estimate** | **se** | **Wald Z** | **p-value** | **95% CI** |
| Between variance | 211.926 | 16.191 | 13.706 | < .001 | [192.356 – 256.041] |
| Within variance | 69.907 | 5.984 | 11.862 | < .001 | [59.110 – 82.677] |
| Intraclass correlation | 0.752 | - | - | - | - |

**Table 7g.** Results of the test-retest analysis: mixed models for CPHQ health literacy

| Effect | Estimate | se | t | p-value | 95% CI |
|---|---|---|---|---|---|
| **Intercept** | 80.363 | 0.732 | 109.763 | < .001 | [78.926 – 81.801] |
| **Time [ref = T0]** | | | | | |
| T1 | -0.714 | 0.460 | -1.551 | 0.122 | [-1.618 – 0.190] |
| T2 | -0.391 | 0.462 | -0.848 | 0.397 | [-1.297 – 0.514] |
| **Adverse events [ref = No]** | | | | | |
| Event at T0 | 0.423 | 0.946 | 0.447 | 0.655 | [-1.435 – 2.280] |
| Event at T1 | 0.531 | 1.265 | 0.420 | 0.675 | [-1.954 – 3.015] |
| Event at T2 | -0.106 | 1.228 | -0.086 | 0.932 | [-2.518 – 2.307] |
| **Positive events [ref = No]** | | | | | |
| Event at T0 | 1.506 | 1.027 | 1.467 | 0.143 | [-0.511 – 3.522] |
| Event at T1 | -1.931 | 2.302 | -0.839 | 0.402 | [-6.453 – 2.591] |
| Event at T2 | -0.253 | 3.291 | -0.077 | 0.939 | [-6.718 – 6.212] |
| **Covariance parameters** | **Estimate** | **se** | **Wald Z** | **p-value** | **95% CI** |
| Between variance | 92.485 | 7.228 | 12.796 | < .001 | [79.351 – 107.794] |
| Within variance | 57.509 | 3.381 | 17.010 | < .001 | [51.250 – 64.532] |
| Intraclass correlation | 0.617 | - | - | - | - |

**Table 7h.** Results of the test-retest analysis: mixed models for CPHQ autonomy

| Effect | Estimate | se | t | p-value | 95% CI |
|---|---|---|---|---|---|
| **Intercept** | 77.870 | 0.863 | 90.196 | < .001 | [76.175 – 79.565] |
| **Time [ref = T0]** | | | | | |
| T1 | -0.322 | 0.456 | -0.707 | 0.480 | [-1.217 – 0.573] |
| T2 | -0.691 | 0.467 | -1.479 | 0.139 | [-1.607 – 0.226] |
| **Adverse events [ref = No]** | | | | | |
| Event at T0 | -1.958 | 1.138 | -1.720 | 0.086 | [-4.193 – 0.278] |
| Event at T1 | -0.487 | 1.522 | -0.320 | 0.749 | [-3.478 – 2.503] |
| Event at T2 | -1.668 | 1.478 | -1.129 | 0.259 | [-4.572 – 1.235] |
| **Positive events [ref = No]** | | | | | |
| Event at T0 | 0.670 | 1.236 | 0.543 | 0.588 | [-1.757 – 3.097] |
| Event at T1 | 1.636 | 2.770 | 0.591 | 0.555 | [-3.806 – 7.079] |
| Event at T2 | 4.772 | 3.961 | 1.205 | 0.229 | [-3.009 – 12.553] |
| **Covariance parameters** | **Estimate** | **se** | **Wald Z** | **p-value** | **95% CI** |
| Between variance | 143.864 | 10.067 | 14.290 | < .001 | [125.426 – 165.012] |
| Within variance | 56.503 | 3.126 | 18.073 | < .001 | [50.696 – 62.975] |
| Intraclass correlation | 0.718 | - | - | - | - |

**Table 7i.** Results of the test-retest analysis: mixed models for CPHQ total score

| Effect | Estimate | se | t | p-value | 95% CI |
|---|---|---|---|---|---|
| **Intercept** | 76.348 | 0.713 | 107.096 | < .001 | [74.947 – 77.748] |
| **Time [ref = T0]** | | | | | |
| T1 | -0.431 | 0.240 | -1.796 | 0.073 | [-0.903 – 0.040] |
| T2 | -0.457 | 0.234 | -1.956 | 0.051 | [-0.916 – 0.002] |
| **Adverse events [ref = No]** | | | | | |
| Event at T0 | -2.496 | 0.970 | -2.573 | 0.010 | [-4.402 – -0.590] |
| Event at T1 | -1.300 | 1.298 | -1.002 | 0.317 | [-3.849 – 1.249] |
| Event at T2 | -2.483 | 1.260 | -1.971 | 0.049 | [-4.958 – -0.008] |
| **Positive events [ref = No]** | | | | | |
| Event at T0 | 0.944 | 1.053 | 0.896 | 0.371 | [-1.125 – 3.013] |
| Event at T1 | 1.002 | 2.362 | 0.424 | 0.671 | [-3.637 – 5.642] |
| Event at T2 | 2.260 | 3.377 | 0.669 | 0.504 | [-4.373 – 8.894] |
| **Covariance parameters** | **Estimate** | **se** | **Wald Z** | **p-value** | **95% CI** |
| Between variance | 111.754 | 7.212 | 15.496 | < .001 | [98.476 – 126.852] |
| Within variance | 15.697 | 1.030 | 15.233 | < .001 | [13.802 – 17.852] |
| Intraclass correlation | 0.877 | - | - | - | - |

**Table 8.** Results of the test-retest analyses: weighted Kappa analyses (n=543)

| | **T0 vs T1** | | | **T0 vs T2** | | |
|---|---|---|---|---|---|---|
| **Domain** | **Kappa [95% CI]** | **Z** | **p-value** | **Kappa [95% CI]** | **Z** | **p-value** |
| Mental relaxation | 0.759 [0.714-0.804] | 17.690 | <0.001 | 0.709[0.661-0.758] | 16.533 | <0.001 |
| Enjoyment | 0.767 [0.722-0.812] | 17.879 | <0.001 | 0.802 [0.765-0.839] | 18.683 | <0.001 |
| Autonomy | 0.671 [0.616-0.726] | 15.688 | <0.001 | 0.667 [0.612-0.721] | 15.546 | <0.001 |
| Fitness | 0.810 [0.774-0.845] | 18.885 | <0.001 | 0.752 [0.706-0.798] | 17.586 | <0.001 |
| Social acceptance | 0.692 [0.644-0.740] | 16.166 | <0.001 | 0.692 [0.642-0.742] | 16.124 | <0.001 |
| Social support | 0.721 [0.671-0.771] | 16.815 | <0.001 | 0.611 [0.557-0.666] | 14.788 | <0.001 |
| Financial resources | 0.731 [0.679-0.783] | 17.124 | <0.001 | 0.686 [0.629-0.742] | 16.105 | <0.001 |
| Health literacy | 0.621 [0.563-0.679] | 14.495 | <0.001 | 0.603 [0.544-0.662] | 14.105 | <0.001 |
| Total score | 0.795 [0.758-0.831] | 18.541 | <0.001 | 0.790 [0.755-0.825] | 18.433 | <0.001 |
